# Multimodal imaging improves brain age prediction and reveals distinct abnormalities in patients with psychiatric and neurological disorders

**DOI:** 10.1101/2020.06.29.20142810

**Authors:** Jaroslav Rokicki, Thomas Wolfers, Wibeke Nordhøy, Natalia Tesli, Daniel S. Quintana, Dag Alnæs, Genevieve Richard, Ann-Marie G. de Lange, Martina J. Lund, Linn Norbom, Ingrid Agartz, Ingrid Melle, Terje Nærland, Geir Selbæk, Karin Persson, Jan Egil Nordvik, Emanuel Schwarz, Ole A. Andreassen, Tobias Kaufmann, Lars T. Westlye

## Abstract

**Background:** The deviation between chronological age and age predicted using brain MRI is a putative marker of brain health and disease-related deterioration. Age prediction based on structural MRI data shows high accuracy and sensitivity to common brain disorders. However, brain aging is complex and heterogenous, both in terms of individual differences and the biological processes involved. Here, we implemented a multimodal age prediction approach and tested the predictive value across patients with a range of disorders with distinct etiologies and clinical features.

**Methods:** We implemented a multimodal model to estimate brain age using different combinations of cortical area, thickness and sub-cortical volumes, cortical and subcortical T1/T2-weighted ratios, and cerebral blood flow (CBF) calculated from functional arterial spin labeling (ASL) data. For each of the 11 models we assessed the age prediction accuracy in HC n=761 and compared the resulting brain age gaps (BAGs) between each clinical group and age-matched subsets of HC in patients with Alzheimer’s disease (AD, n=54), mild cognitive impairment (MCI, n=88), subjective cognitive impairment (SCI, n=55), schizophrenia (SZ, n=156), bipolar disorder (BD, n=136), autism spectrum disorder (ASD, n=28).

**Results:** Among the 11 models, we found highest age prediction accuracy in HC when integrating all modalities (mean absolute error=6.5 years). Beyond this global BAG, the area under the curve for the receiver-operating characteristics based on two-group case-control classifications showed strongest effects for AD and ASD in global T1-weighted BAG (T1w-BAG), while MCI, SCI, BD and SZ showed strongest effects in CBF-based BAGs.

**Conclusions:** Combining multiple MRI modalities improves brain age prediction and reveals distinct deviations in patients with psychiatric and neurological disorders. The multimodal BAG was most accurate in predicting age in HC, while group differences between patients and controls were often larger for BAGs based on single modalities. These findings demonstrate that multidimensional phenotyping provides a mapping of overlapping and distinct pathophysiology in common disorders of the brain, and specifically suggest metabolic and neurovascular aberrations in SZ and at-risk and early stage dementia.

## Introduction

Brain age gap (BAG) - the difference between an individual’s chronological and predicted age based on imaging data may serve as a surrogate marker of general brain health and disease-related deterioration of the brain (Cole and Franke 2017). Previous studies have focused on morphometric and volumetric features derived from T1-weighted (T1w) images. While this single-modality approach has the benefit of reducing analytic complexity, it does not take into consideration the vast biological heterogeneity of the developing and aging brain, and comes at the cost of reduced sensitivity to biological processes that are not primarily reflected in gross brain anatomy or morphology (Richard et al., 2018).

Normative brain development and aging, as well as emerging disease mechanisms in common brain and mental disorders, are highly heterogenous both in terms of spatial distribution and underlying neurobiology. As brain disorders disrupt brain structure and function on different levels by divergent pathophysiological mechanisms, it is reasonable to assume that such variety cannot be fully captured by a brain estimate derived from a single MRI modality. Therefore, single-modality approaches may diminish sensitivity and specificity to clinical conditions with distinct pathophysiology, and interpreting clinical brain aberrations based solely on T1w-data may conceal relevant information. In fact, various brain imaging modalities reveal unique patterns and trajectories across the lifespan, and possibly capture distinct biological contributions to brain aging and disease development. While the large majority of brain age prediction studies have focused on T1w data only, some studies have simultaneously considered numerous imaging modalities in order to model multiple biologically distinct brain ages (Smith et al., 2020, Liem et al., 2017, Richard et al., 2018, Brown et al., 2012, Cole, 2020, Engemann et al., 2020, Niu et al., 2020). However, few attempts have been made to model multiple biologically distinct brain ages in patient populations. In addition to overall increased age prediction accuracy offered by the complementary information, considering a range of MRI modalities may provide more insight into specific deleterious neurobiological processes associated with different clinical conditions.

A number of brain disorders have been associated with increased BAG, with schizophrenia (SZ) and Alzheimer’s disease (AD) among those showing largest effects, primarily driven by apparent cortical thinning and aberrant subcortical volumes (Kaufmann et al. 2018). BAG has been shown to be more accurate in predicting the conversion of mild cognitive impairment (MCI) to AD compared to T1-based MRI features such as cortical thickness and regional brain volumes (Gaser 2013) and potentially can be used as a biomarker for early dementia risk screening (Wang et al., 2019). Bipolar disorder (BD) and schizophrenia (SZ) are severe mental disorders with overlapping symptoms and shared neurobiological underpinnings (Ruderfer et al. 2018). Whereas previous imaging studies aiming at identifying brain morphometric correlates of the disorders have revealed diverging results, differences in BAG between SZ and HC were shown to be larger than differences between BD and HC (Kaufmann et al. 2018, Hajek et al. 2017). Autism spectrum disorders (ASD) are neurodevelopmental disorders affecting social and communication skills, and exhibit overlapping pathophysiological features and genetic underpinnings with SZ (Foss-Feig, 2019, Lee et al., 2013). A recent study linked higher BAG in youth with ASD to lower disease severity (Tunç et al. 2019).

Several structural imaging modalities and markers have been proposed. Myelin deficiencies and abnormal expression of myelin genes are common in brain disorders, and, while conflicting evidence exists (Stokowy et al., 2018), previous reports have implicated oligodendroglial dysfunction and abnormalities in myelin maintenance and repair in SZ (Davis et al., 2003, Jungerius et al., 2008, Lee and Fields, 2009). Previous studies have demonstrated that intra-cortical myelin content, as assessed using T1/T2-weighted (T1w/T2w) ratio, shows a characteristic pattern of aging-related effects (Grydeland et al. 2013a, Grydeland et al., 2018) and is affected in both SZ (Iwatani et al. 2015; Ganzetti, Wenderoth, and Mantini 2015) and BD (Ishida et al. 2017). Furthermore, associations between risk of developing psychosis, cognitive ability and apparent myelin content were found in children and adolescents (Norbom et al. 2019). An influential myelin model suggests a close relationship between degenerative disorders such as AD and myelination (Bartzokis, 2011), yet, recent studies using T1w/T2w ratio revealed contradicting results, reporting both cortical demyelination (Luo et al., 2019) and higher cortical myelin content (Pelkmans et al., 2019) in AD.

While the T1w/T2w ratio partly reflects the cortical myeloarchitecture, Arterial spin labeling (ASL) enables a non-invasive assessment of cerebral blood flow (CBF), serving as a proxy for clinically relevant neurovascular and metabolic processes in the brain (Haller et al. 2016, Biagi et al. 2007). Accordingly, recent studies reported case-control differences in CBF in individuals with SZ (Stegmayer et al. 2017a, Pinkham et al. 2011), sub-clinical psychotic-type experiences (Modinos et al. 2018), ASD (Ota Miho, 2018, Yerys et al., 2018), and in various forms and severities of neurodegenerative disorders (Zhang et al., 2017).

In this study, we tested for dissociable brain deviations in common neurological and psychiatric disorders based on BAG defined using measures of brain morphology (cortical thickness, cortical area, subcortical volumes), T1w/T2w ratio maps partially reflecting cortical and subcortical myelin content, and cortical and subcortical CBF maps reflecting neurovascular and metabolic processes in the brain. We used data from HC as training sets and for comparison of age prediction accuracy across modalities. We included data from patients with aging-related cognitive and neurodegenerative disorders, including subjective cognitive impairment (SCI), MCI and AD, as well as severe mental disorders with a neurodevelopmental etiology, including SZ, BD and ASD. All subjects were scanned on the same scanner with a standardized imaging protocol. Based on the assumption that the different imaging modalities provide complementary information of brain structure and function, we hypothesized that the model fusing all modalities would show highest age prediction accuracy. Based on previous studies, we anticipated overall abnormalities in BAG across disorders, yet given the differential etiology and pathophysiology of the neurodegenerative and neurodevelopmental spectrum disorders, we expected that the single-modality BAGs would show differential sensitivity in the distinct groups.

## Methods

Figure 1 shows an overview of the general approach.

**Figure 1.**
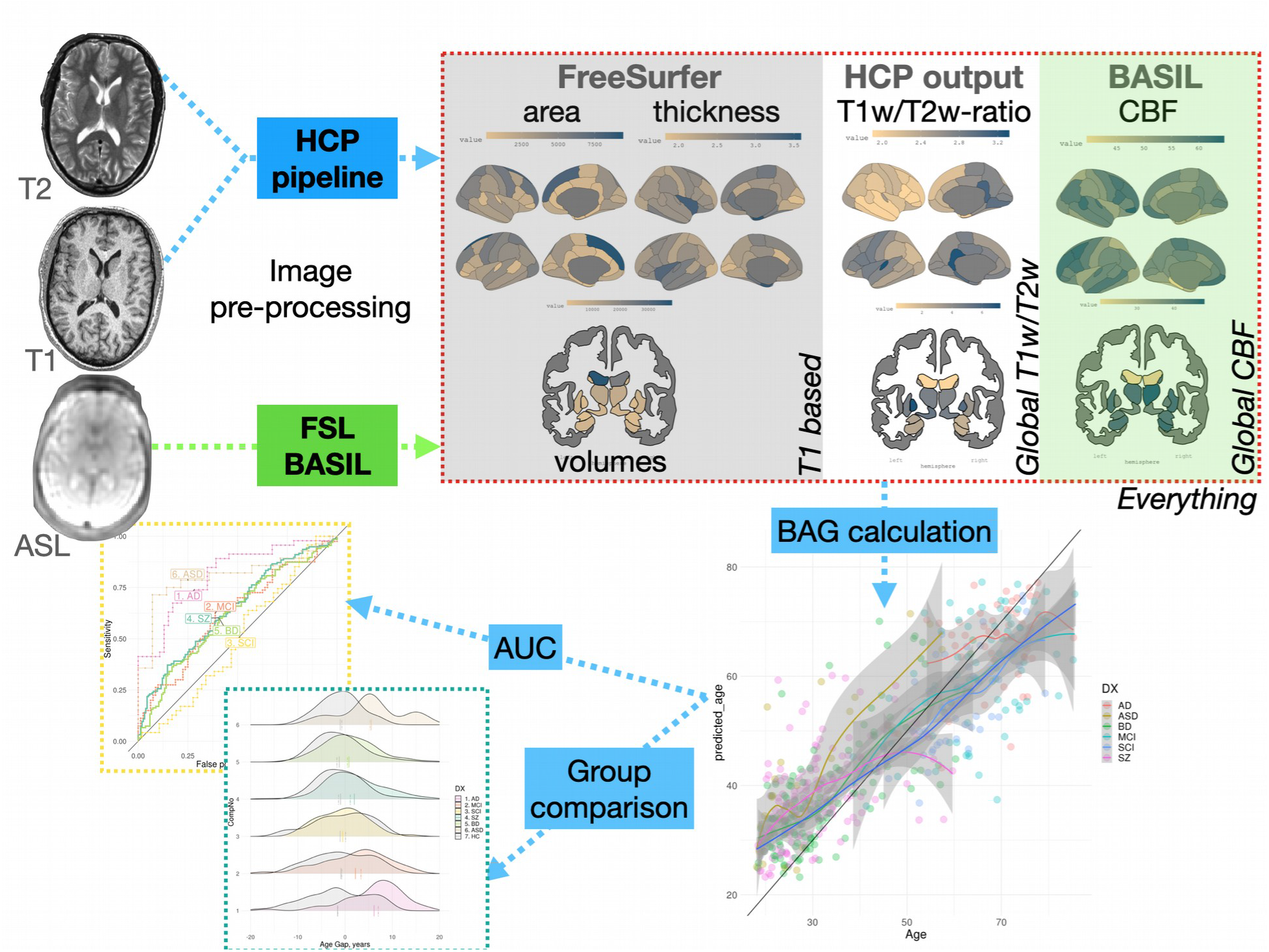
Pipeline. Three MRI sequences were used to generate eleven feature modalities to build brain age prediction models. These models trained using HC subjects to predict age. Then, we applied these models to patient populations. Subsequently, group tests were performed building a linear model for each patient group age and sex matched with controls.

### Participants

T1w, T2w and ASL data were available from 1546 participants. Among these, 268 were excluded due to insufficient T1w image quality (n=139), errors occurring during the Freesurfer pipeline (n=7), errors in estimation of T1w/T2w ratio maps (n=42), errors in CBF calculations (n=18), missing demographic information (n=2) and unspecific or unknown clinical diagnosis (n=60). The final sample thus consisted of data from 1278 individuals. All participants were recruited from the Greater Oslo Region as part of four studies: the Thematically Organized Psychosis (TOP) study, the Norwegian register of persons assessed for cognitive symptoms (NorCog), the BUPGEN study on ASD and other neurodevelopmental disorders, and STROKEMRI study (Richard et al., 2018). Key demographics are described in details in Table 1, and Supplementary Figure S1 and Table S1. Clinical information is summarized in Supplementary Table S2.

**Table 1.** Participant demographics summarized by diagnosis (N - number of participants, SD – standard deviation)

| **GROUP** | **N** | **MEAN AGE,**<br>**YEARS** | **SD AGE** | **MIN**<br>**AGE** | **MAX**<br>**AGE** | **% MALE** |
| --- | --- | --- | --- | --- | --- | --- |
| HC | 761 | 44.9 | 16.1 | 18.0 | 85.8 | 56.0 |
| AD | 54 | 69.4 | 6.8 | 54.2 | 85.3 | 38.9 |
| MCI | 88 | 65.8 | 9.7 | 38.6 | 85.5 | 55.7 |
| SCI | 55 | 59.3 | 8.7 | 39.6 | 79.3 | 41.8 |
| SZ | 156 | 31.1 | 9.2 | 18.4 | 59.7 | 53.8 |
| BD | 136 | 32.7 | 10.8 | 18.4 | 63.9 | 36.0 |
| ASD | 28 | 29.6 | 11.9 | 18.1 | 57.4 | 64.3 |
| **TOTAL** | **1278** | **44.7** | **17.3** | **18.0** | **85.8** | **52.4** |

Inclusion criteria for participants in TOP were meeting the DSM-IV diagnostic criteria for broad schizophrenia and bipolar spectrum diagnoses, age between 18 and 65 years, no history of severe head injury or other disorders affecting the central nervous system, and an IQ>70. The participants were referred to the study by their clinicians from local hospitals. HC were recruited through a stratified random selection from national records. Upon inclusion in the TOP study, the HC were screened with the Primary Care Evaluation of Mental Disorders (PRIME-MD) in order to confirm no history of psychiatric disorder.

NorCog participants were recruited from the memory clinic at Oslo University Hospital. Diagnosis of AD was based on ICD-10 criteria, while MCI was defined according to Winblad criteria (Winblad et al. 2004). Patients with subjective cognitive disturbances who did not meet the Winblad criteria were also included in the study as SCI. The diagnoses were confirmed by experienced specialists in geriatric medicine, neurology or psychiatry.

Patients diagnosed with ASD were sampled from the BUPGEN study, which recruits patients from Norwegian health services specializing in assessment of ASD and other neurodevelopmental disorders. The main BUPGEN inclusion criterion was that either a community service or the specialized health services had referred the patient to a mental health center for a diagnostic assessment of ASD. The patients were assessed by experienced clinical psychologists and psychiatrists. Diagnostic conclusions were best-estimate clinical diagnoses derived from tests, interview results and observations. All diagnoses were based on the International Statistical Classification of Diseases and Related Health Problems 10th Revision (ICD-10) (Reindal et al., 2019).

HC in STROKEMRI were recruited through local newspapers and social media, were required to be at least 18 years old and reported no history of stroke, dementia, or other neurological and psychiatric disorders, alcohol and substance abuse or intake of medications significantly affecting the nervous system.

The studies were approved by the Regional Committee for Medical Research Ethics and the Norwegian Data Inspectorate and written informed consent was obtained from all participants.

### MRI acquisition

MRI data was collected on a 3 T DiscoveryTM (MR750) scanner (GE Healthcare, Milwaukee, WI, US) located at the Oslo University Hospital equipped with the vendor’s 32-channel head coil. Whole brain isotropic T1w structural data was acquired using an inversion recovery-fast spoiled gradient echo sequence (BRAVO) with the following parameters: TR=8.16 ms, TE=3.18 ms, TI=450 ms, flip angle=12°, field of view=256 mm, acquisition matrices=256×256, 188 sagittal slices, slice thickness=1.0 mm, voxel size=1×1×1 mm^3^. Whole brain isotropic T2w structural image was acquired using a 3D fast spin echo (FSE) sequence (CUBE) with following parameters: TR=2500 ms, TE=7.5 ms, field of view=256 mm, acquisition matrices=256×256, 188 sagittal slices, slice thickness=1 mm, voxel size=1×1×1 mm3. The ASL scan being a pseudo-Continuous ASL (pCASL) with a 3D FSE interleaved stack-of-spirals (3D spiral) readout with 512 points on 8 spirals and an acquisition resolution of 4×4×3.0 mm3, reconstructed by default to a 2×2×3 mm3, TE=11.072 ms, TR=5025 ms, Labeling duration (LD)=1450 ms, post labeling Delay (PLD)=2025 ms, number of excitations (NEX)=3. The PLD was chosen as recommended by the review article of Alsop et al. (Alsop et al, MRM, 2015) for clinical patients. All the scans were performed in the same session.

### Image quality control

Image quality control was performed as a two-step process. First all images were processed with MRIQC (Esteban 2017). The T1w images classified by a default machine learning algorithm to an exclude node with a probability of at least 0.6 were removed from subsequent analyses. In the second step, all the final cortical maps (area, T1w/T2w ratio and CBF) were visually quality checked.

### Image pre-processing

Cortical thickness and T1w/T2w ratio maps were created using Human Connectome Project (HCP) pipeline based on *Freesurfer v6*.*0* (Fischl, 2012). Briefly, the T2w images were co-registered to the T1w images, followed by registration to the MNI space. Subsequently, cortical surfaces were extracted using *Freesurfer* and used for estimation of cortical thickness and area. We modified the original *Freesurfer* pipeline by removing -t2pial flag after careful considerations of quality control.

CBF maps were estimated using *Bayesian Inference for Arterial Spin Labeling MRI (BASIL)* (Chappell et al. 2009) in FSL (Jenkinson et al., 2012). When processing the ASL data such parameters were used (tissue T1=1.2 s, arterial T1=1.6 s, labeling efficiency=0.6, bolus arrival time = 1.45 s, bolus duration = 1.45 s, inversion time TI = 3.475 s, delay between the end of labeling and the start of the acquisition 2.025 s and blood-brain barrier coefficient 0.98 ml/g). Equilibrium blood magnetisation was estimated voxel-wise by using the M0 image. The CBF map was spatially smoothed using an adaptive filter, which exploited neighboring voxel signals on an intensity-dependent basis, without interacting with non-linear kinetics where smoothing was unnecessary (Groves, Chappell, and Woolrich 2009). Due to low spatial resolution of the perfusion image, partial volume error correction was performed to improve the accuracy of CBF estimation (Chappell et al. 2011). CBF maps were mapped to the individual cortical surface from *Freesurfer* for further analysis.

For between-subject analysis we extracted values for cortical thickness and area, CBF and T1w/T2w surface ratio from 34 cortical regions of interest (ROIs) in each hemisphere based on the Desikan-Killiany atlas (Desikan et al., 2006). In addition, we computed the mean CBF and T1w/T2w ratio within 35 subcortical regions based on the automated volume segmentation in *Freesurfer* (Supplementary material 3).

### Brain age gap calculation

For brain age prediction we subdivided our measures in four cortical feature sets: thickness, area, T1w/T2w ratio and CBF, and three subcortical sets: volumes, T1w/T2w ratio and CBF. We also integrated all T1w image derived measures (area, thickness and volume) into one composite set comprising 171 features, referred as “global-T1w”. The same procedure was repeated for cortical and subcortical T1w/T2w ratio and CBF. This yielded a set of 103 features for each modality. Finally, we gathered data from all modalities to a single set with 377 features, referred as “global-multimodal”. We investigated 11 feature sets in total.

Brain age of each individual was calculated by growing 5000 trees using *random Forest package* in R (Breiman 2001), which was chosen for both its resilience to overfitting and its robustness to noise (Breiman 2001). First, we divided our sample into HC and patients. For HC, we used 10-fold cross validation, whereas for the patients the model was trained on all the HC data and subsequently applied to the individual patients. This approach was chosen to keep the relatively few HCs aged 75 years and older (n=23) in the training set in order to retain predictive power at older ages. Cortical thickness, T1w/T2w ratio and CBF features were residualized for sex using linear models, whereas subcortical volumes and cortical areas were residualized for both sex and intracortical volume (ICV). To account for a well-known bias in brain age prediction, we residualized brain age with respect to age using linear models (G de Lange and Cole, 2020) (Figs. S16-S17). Furthermore, to assess their complementary value we estimated Spearman’s correlation between BAGs derived from different modalities in HC and applied hierarchical clustering based on Ward’s minimum variance criterion (Ward, 1963), as implemented in *corrplot* R package (Wei and Simko 2017).

We report root mean squared error (RMSE), mean absolute error (MAE) and shared variance (r^2^) between predicted and chronological age as measures of prediction accuracy for all models. In order to estimate the features contributing the most to brain age prediction we used mean increase in mean squared error (MSE). This measure quantifies the difference in MSE between randomly shuffled and actual values for the investigated feature, while keeping the rest of the features intact when applied on unseen data. To visualize feature importance MSE was mapped onto segmented brain surface using *ggseg* R package (Mowinckel and Vidal-Pieiro 2019).

### Group comparison

In order to assess group differences between patients and controls we matched subjects to controls with respect to age and sex using nearest neigbor matching with 1:1 ratio and logistic regression distance as implemented in the R package *matchIt* (Ho et al. 2007). Demographics for each group comparison are listed in Table S3. Subsequent group tests were performed building a linear model for each case control pair while controlling for age and sex.To correct for multiple tests (6 disorders × 11 modalities, 66 in total), reported p-values were adjusted using a false discovery rate (FDR) threshold. Additionally, to assess effect sizes we converted t-statistics to Cohen’s d. Lastly, for each derived BAG we calculated the area under the curve (AUC) of receiver operating characteristics (ROC) in order to evaluate its performance in a two-class classification setting as implemented in R *metrics* package. Furthermore, we computed sensitivity and specificity at the optimal cut-off point. To estimate significance of AUC we performed 5000 permutations with random shuffling of the group label to build null distributions. The associated p-value based on the empirical null distribution was adjusted for number of modalities and patient groups using FDR. A two-class design was chosen due to modest overlap in age between distinct patient groups.

## Results

### Predicting brain age in healthy controls

To train the model we used data obtained from 761 healthy individuals. The model integrating all modalities and feature sets showed the best fit with r^2^ = 0.77 (MAE=6.5 years), with global T1w/ T2w ratio and global T1w derived features having very similar fits: r^2^ = 0.73 (MAE=6.9) and r^2^ = 0.72 (MAE=6.9), respectively (Fig. 2a). Clustering based on Ward’s criterion (Fig. 2b) suggested 4 main BAG clusters: 1) CBF modalities, 2) cortical thickness with cortical T1w/T2w ratio, 3) cortical area, and 4) subcortical T1w/T2w ratio, subcortical volumes, global T1w/T2w ratio, global T1w based and global multimodal. Fit for other modalities and feature sets with corresponding MAE are listed in Supplementary Table S4. Mean values for each modality were summarized by decades (Figs. S2-S8) as well trajectories of mean cortical (Figs. S9-S12) and hippocampal (S13-S15) values.

**Figure 2.**
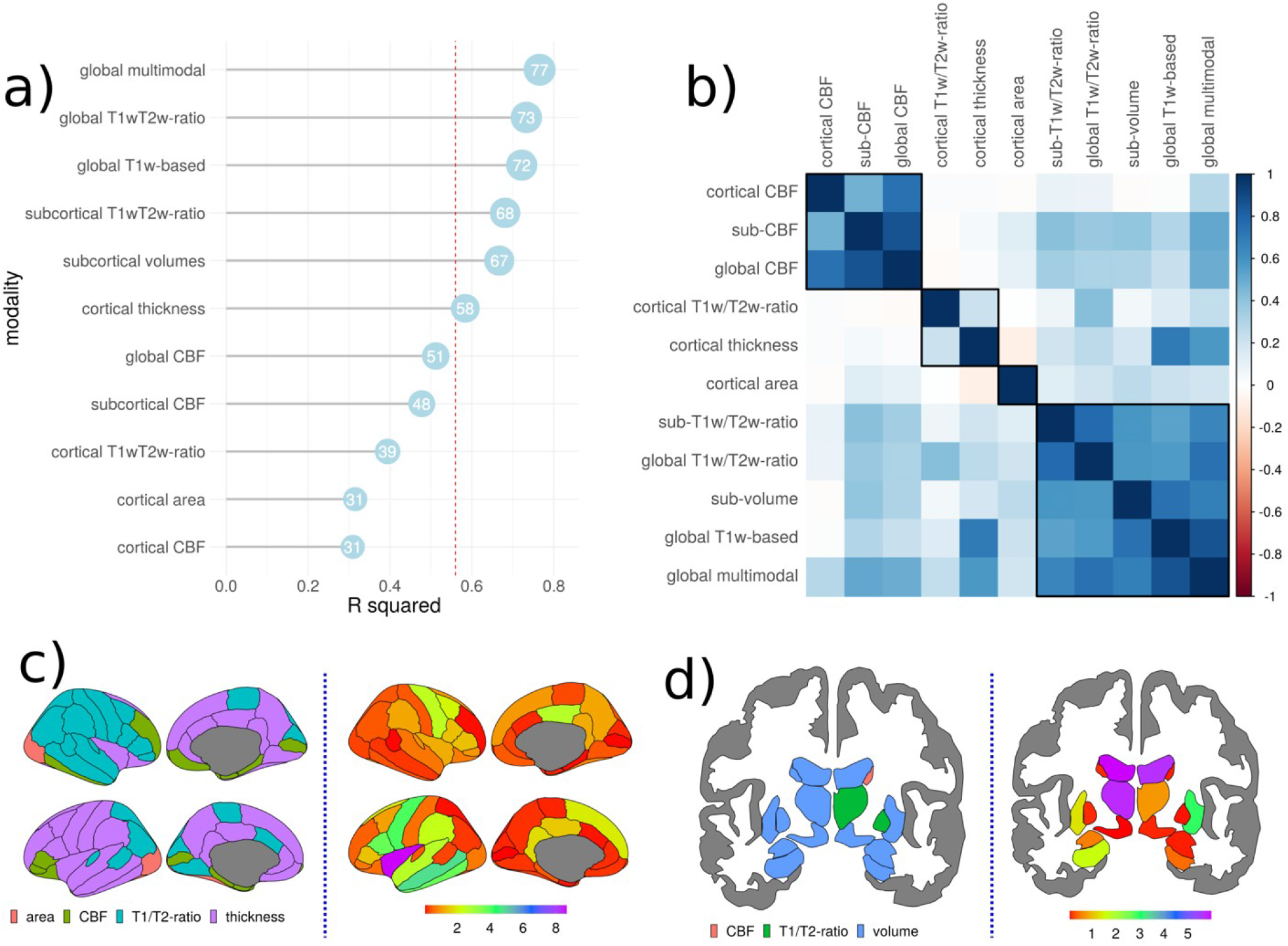
HC model fit. BAGs ranked from the most (top) to the least (bottom) accurate based on out of sample r^2^ (multiplied by 100) of the model (a) and BAG Spearman’s correlation matrix with 4 clusters marked by black lines (b). Modality from which a given feature was derived (left) and feature importance measured as increase of MSE (right) shown as a colormap overlaid on the brain for the best model integrating all modalities in cortex (c) and in subcortical structures (d).

The features contributing most to age prediction in the model comprising all features were predominantly derived from cortical thickness, subcortical volumes and T1w/T2w ratio (Fig. 2c). The three most contributing features in cortex were thickness in insula, middle temporal and precentral and gyri in the left hemisphere. Among the subcortical structures, the strongest features were T1w/T2w ratio and volume of 3^rd^ ventricle, followed by right lateral ventricle volume and T1w/T2w ratio and thalamus volume (Fig. 2d). Among the 20 most predictive features 11 were subcortical (3 T1w/T2w ratio and 8 subcortical volumes), and 9 were cortical (8 thickness and one T1w/T2w ratio based). The top 20 features with importance estimates for the best model are given in Supplementary Table S5. Feature importance for other modalities is presented in Supplementary Figures S18 and S19.

### Group differences in BAG

In the second step we applied the models trained on HC to the clinical samples and performed case-control comparison of the resulting BAGs. The results are shown in Figure 3 and Supplementary Figure S20. All modalities apart from the one based on T1w/T2w ratio showed significantly higher BAG in AD compared to HC, with global T1w-BAG exhibiting largest effect (7.4 years, p<.001, Cohen’s d=1.09), followed by subcortical volumes (7.9 years, p<.001, d=1.04) and cortical thickness (6.9 years, p<.001, d=0.96). BAG based on global T1w/T2w ratio showed group differences in patients with AD (p<.05, d=0.47), SZ (p<.05, d=0.30), and ASD (p<.05, d=0.63), while having the second best fit in brain age prediction in HC population.

**Figure 3.**
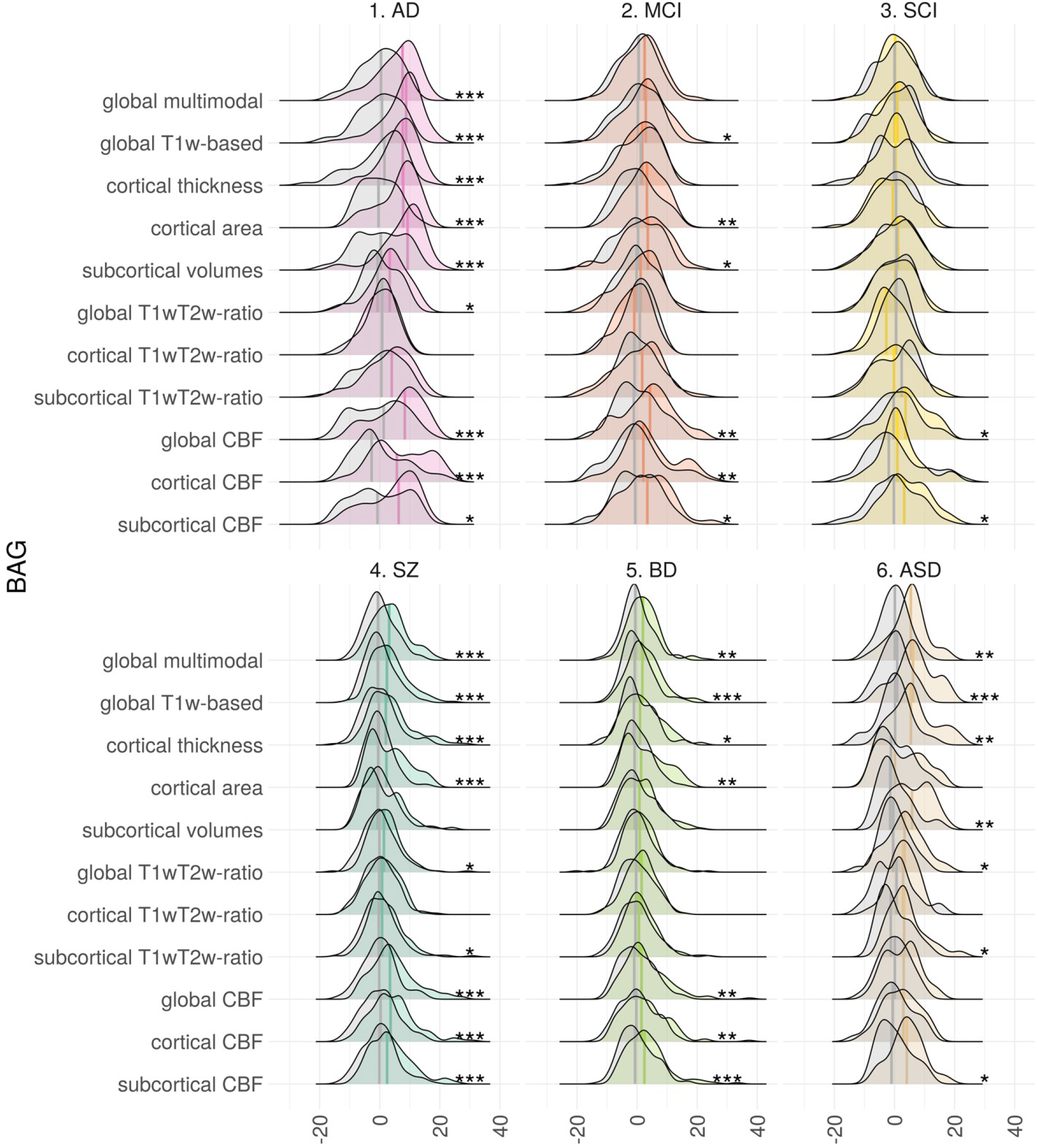
Group comparison of BAG in HC vs patient groups. Both distributions and means are shown. Stars on the right side indicate significant results (FDR corrected for multiple comparisons), with p<0.05, p<0.01 and p<0.001 being marked as 1 to 3 stars. Distributions for HC are show in gray.

Across modalities, mean BAG for AD was on average 5.3 years higher than for HC. For MCI and SCI the difference was 2.0 and 0.8 years, respectively. For MCI and SCI the largest differences from HC were seen in CBF-based BAGs. For individuals with MCI cortical-CBF BAG was 4.0 years higher than HC (p<.01, d=0.45), followed by global CBF (3.7 years, p<.01, d=0.47). For individuals with SCI largest BAG deviations from matched HC were based on subcortical CBF with 4.1 years (p<.05, d=0.54), followed by global CBF (3.6 years, p<.05, d=0.47). These were the only two BAGs showing significant case-control differences for SCI.

BAG was significantly higher in SZ compared to HC for almost all modalities. The strongest group differences were found for global CBF-BAG (4.5 years, p<.001, d=0.68), followed by cortical CBF (4.2 years, p<.001, d=0.61). Apart from the CBF models, the global multimodal model yielded similar effect size as cortical CBF (3.5 years, p<.001, d=0.65). Across modalities, the average BAG in SZ was 2.7 years higher than HC.

For BD, subcortical CBF-BAG showed largest effect size (3.0 years, p<.001, d=0.48), followed by global CBF-BAG (2.8 years, p<.01, d=0.42), and T1w-based BAG (2.5 years, p<.001, d=0.48). Across modalities, average BAG in BD was 2.0 years higher than HC.

BAG derived from global T1w features showed strongest group differences between patients with ASD and HC (6.7 years, p<.001, d=1.19), followed by cortical thickness BAG (6.5 years, p<.01, d=1.00). Across modalities, average BAG in BD was 4.1 years higher than HC.

Complete list of results is summarized in Supplementary Table S6.

### Group differences in BAG

To evaluate classification accuracy of each modality we calculated AUC of ROC for pairwise group classifications of cases and controls for each clinical group. Then, we built a null distribution using 5000 random permutations to estimate p-value associated with AUC and FDR corrected it for number of modalities and patient groups. Figure 4 summarizes the main results, and Supplementary Tables S7-S9 provide a full list of AUC for each modality together with sensitivity and specificity at optimal cut-off point. Briefly, we found that for AD global-T1w measures were the most accurate (AUC=0.81, p<.001) followed by subcortical volumes (AUC=0.77, p<.001). For MCI cortical area (AUC=0.64, p<.01) and cortical CBF-BAG (AUC=0.64, p<.01) showed highest AUC. Lastly, for SCI subcortical CBF-BAG was most discriminative (AUC=0.65, p<.01). In psychiatric disorders, SZ was most distinguishable using BAG based on global multimodal features (AUC=0.68, p<.001) along with global CBF (AUC=0.68, p<.001). BD showed consistently lower accuracy as compared to SZ, with highest AUC scores from global T1w-based, global multimodal and subcortical CBF (all AUC=0.63, all p<.001). Finally, ASD was most discernible using global T1w-features (AUC=0.8, p<.01), followed by global multimodal (AUC=0.78, p<.01) and cortical thickness (AUC=0.75, p<.01).

**Figure 4.**
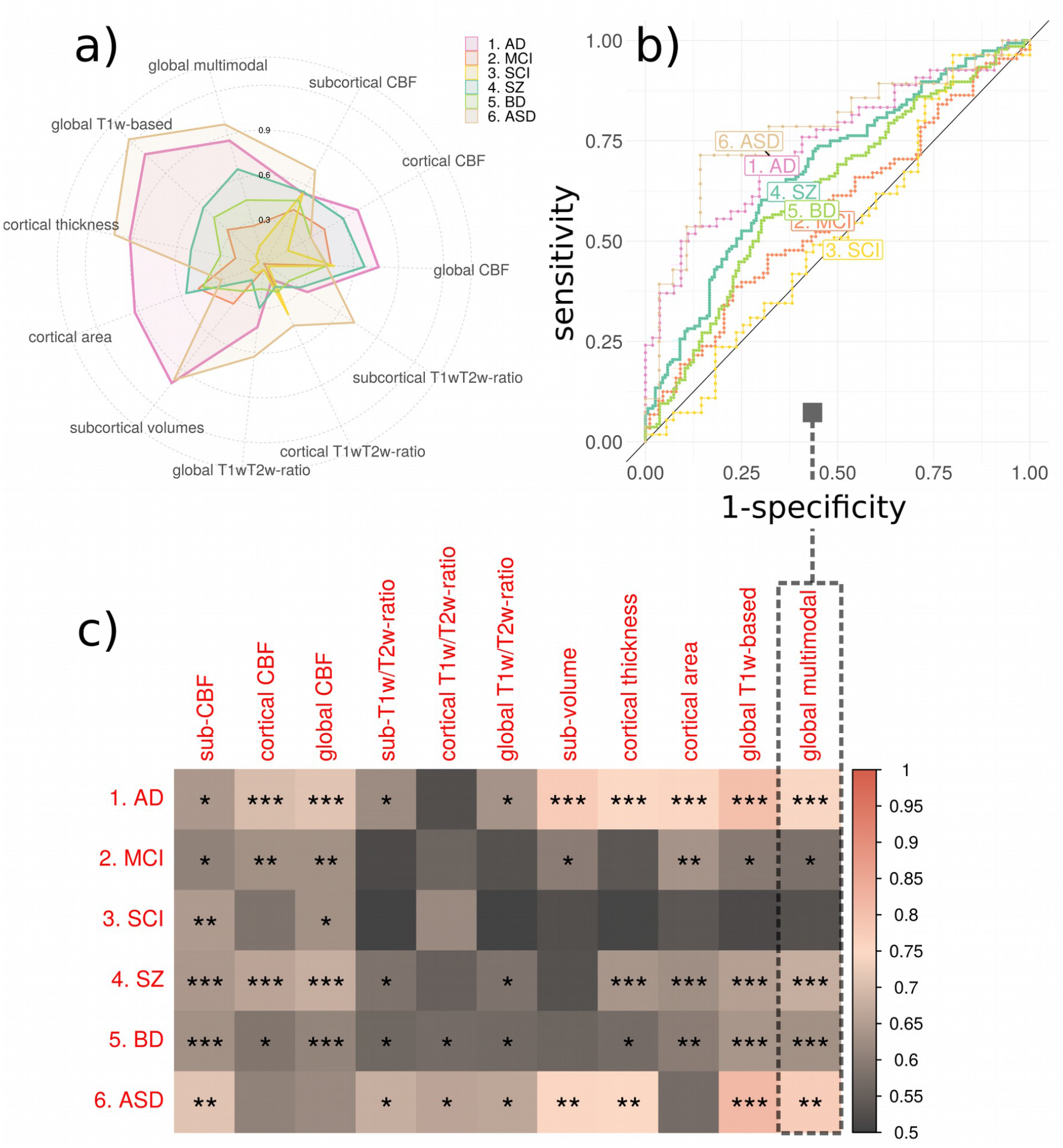
AUC for each group comparison and modality. Spider plot of effect sizes for disorders (a). Receiver operating characteristics (b) are shown for the most accurate model in HC (everything) in the AUC matrix (c)

## Discussion

In this study, we investigated deviations between chronological age and brain age based on multiple brain imaging modalities, including T1w, T2w and ASL data, which all convey distinct biological information. We found high age prediction accuracy for most of the included MRI features in healthy participants, with highest accuracy for the model including all available features. Subsequent case-control comparisons revealed high sensitivity to group differences, with varying performance of the single-modality BAGs, likely corresponding to the distinct underlying neurobiology of each disorder. This demonstrates that brain age aberrations of brain diseases are better characterized and differentiated using multimodal as compared to unimodal imaging approaches.

Highest age prediction accuracy in HC was achieved when integrating all features from all modalities, followed by global T1w/T2w ratio and global T1w-based features. Estimates of feature importance revealed that the model integrating all modalities was mostly driven by the cortical thickness, T1w/T2w ratio and subcortical volumes. The lifespan trajectories derived from these features have been described in several studies. For example, cortical thickness and subcortical volumes decrease monotonically throughout the adult lifespan (Westlye et al., 2010, Fjell et al. 2015, Li et al. 2014). Conversely, the lifespan trajectory of the intra-cortical T1w/T2w ratio has been characterized by a global increase until the end of the 30s, followed by a period of relative stability and subsequent decrease from the end of 6th decade of life (Grydeland et al., 2013a), which is largely in line with our observations. The models with the lowest age prediction accuracy in HC were based on CBF as well as the features derived from cortical area. Previous studies have shown that CBF is highest in children, followed by slow decreases into adolescence until early adulthood, and a period of modest and relatively stable decline until the age of 80 (Biagi et al. 2007, Haller et al., 2016). Such trajectories are in line with our data, and correspond with the moderate performance of CBF for brain age prediction. Additionally, cortical area, while showing a steady age-related decrease starting in the early twenties, is less affected throughout the adult lifespan as compared to cortical thickness or volumes (Storsve et al. 2014).

The prediction performance in the patient populations varied across the different disorders. T1w-based BAGs were most robust in classifying AD and ASD, but were also sensitive to both SZ and BD. In general, the results were in line with a recent large-scale implementation focusing on T1w-based age prediction (Kaufmann et al. 2018) with the exception of ASD, which in our study showed increased BAG. This discrepancy between studies for ASD is likely partly explained by the different age range and clinical characteristics. While Kaufmann *et al*. study primarily included young patients with ASD starting from 5 years of age (mean age 16.4, SD=9.0), we recruited adult patients from habilitation clinics often handling patients with intellectual disability and syndromes. Previous imaging studies on ASD have revealed larger brain volumes during infancy, followed by accelerated decline and degeneration (Courchesne et al., 2011). Moreover, age dependent abnormalities in cortical thinning during childhood, adolescence and early adulthood (Zielinski et al., 2014), and alterations in cerebral blood flow (Ota Miho, 2018) were reported. In addition to the clinical characteristics, the temporal dynamics of key neurodevelopmental processes during childhood and adolescence are possibly contributing to the diverging results between studies (Tunç et al. 2019, Kaufmann et al. 2018). However, caution is warranted when interpreting our particular findings as the current ASD group consisted of only 28 patients.

We identified higher BAG in BD compared to HC using T1w based features. Compared to healthy participants, patients with SZ and BD were shown to have thinner cortex as well as smaller global and regional brain volumes (Boos et al., 2007, Hibar et al., 2016, Hibar et al., 2018), with smaller effects in BD patients (Yao et al., 2017). In line with previous studies (Kaufmann et al., 2018, Hajek et al., 2017) our current analysis supported that T1w based BAG in BD was lower as compared to SZ. Additionally, CBF based features and cortical area showed significantly increased BAG for BD patients. Compared to their healthy peers, patients with BD have also previously been shown to have lower CBF (Toma et al., 2018). In general, our are comparable with classification accuracies reported in other T1w based studies (Nunes et al., 2018, Schnack et al., 2014, Rocha-Rego et al., 2014).

CBF-based BAG also revealed large deviations between HC and SZ. Altered CBF has previously been reported in patients with SZ across a range of brain regions including temporal and parietal lobes, middle frontal gyrus, left putamen as well as superior corona radiata (Stegmayer et al. 2017b, Pinkham et al. 2011). Several of these features, including CBF in the inferior temporal lobe were also among top features in our age prediction model trained on HC. Hence, the deviations in blood flow may index biological processes mimicking pathogenic brain aging in this patient group, contributing to the structural and functional detrimental alterations corroborated by previous studies (Stegmayer et al. 2017a, Pinkham et al. 2011, Ota Miho, 2018).

Assuming that AD reflects a clinical endpoint of chronic deleterious neurodegenerative processes emerging decades prior to symptom onset (Elliott et al. 2019), it is crucial to identify surrogate biomarkers that would be sensitive to early changes when the potential for interventions may be largest. In the neurodegeneration model of AD, the structural brain changes captured by T1w are preceded by metabolic modulations (Jack et al. 2010). Regions with altered metabolism have been linked with altered perfusion in patients with AD (Riederer et al. 2018). Interestingly, BAG based on subcortical CBF had the highest prediction accuracy in SCI, while cortical CBF showed highest case-control difference in MCI, supporting the overall utility of CBF as a sensitive imaging marker of neurovascular and metabolic processes in early phases of dementia. No other modalities revealed significant differences between HC and SCI.

Global T1w/T2w ratio resulted in the second highest age prediction accuracy in HC, but the corresponding BAGs showed only modest case-control differences for the clinical groups. The global model was mostly driven by subcortical T1w/T2w ratio, whereas the cortical T1w/T2w ratio had poorer performance in predicting age in HC. The T1w/T2w ratio intensity measure is assumed to be inversely proportional to cortical myelination and informative for cortical parcellation and structure-function brain mapping (Glasser et al. 2016). However, T1w/T2w ratio is probably not specifically related to myelin content, suggesting a more complex underlying biology (Hagiwara et al. 2018; Ritchie, Pantazatos, and French 2018).

T1w gray-white tissue contrast, yet another measure likely partly associated with cortical myelin content, previously demonstrated high sensitivity to age (Salat et al., 2009, Westlye et al., 2010) and case-control differences have been found in patients with SZ (Jørgensen et al., 2016) and ASD (Mann et al. 2018). It is possible that the same underlying mechanisms explain our findings of higher cortical T1w/T2w ratio BAG in patients with ASD. More specifically, post central gyri previously exhibited one of the largest gray-white tissue contrast reductions in ASD (Mann et al. 2018) and was also among the current top features for age prediction in our sample. We found global T1w/T2w based BAG to be affected in patients with AD and SZ. This is in line with reports of abnormal T1w/T2w in AD (Pelkmans et al. 2019, Grydeland et al., 2013b). Further, emerging evidence indicates lower T1w/T2w in SZ, implicating subcortical (Iwatani et al. 2015; Ganzetti, Wenderoth, and Mantini 2015) and cortical regions (Wei et al., 2020).

It is worth noting some limitations of our study. First, the prediction performance (AUC<0.7) in the patient populations is too low for clinical utility, and was used in the present study to compare the different models. Secondly, our cross-sectional design does not allow us to disentangle the temporal dynamics of the brain aging process, and it remains unclear to which degree the increased BAG in a range of investigated disorders was due to age-related deterioration of the brain or already present since early childhood. Thirdly, we had a modest number of subjects spanning disorders with disparate age ranges, therefore we could not compare them directly. Future studies are also needed to delineate the unique and additive contribution of genetic and environmental variables on the observed group differences. For example, in line with most clinical studies our study design does not allow us to differentiate the effects of disease mechanisms from secondary causes such as lifestyle and medication. This study has several advantages, our study while being cross-sectional covered the whole adult age span. Moreover, all imaging data were obtained on the same scanner using identical sequences and processed using an identical pipeline.

In summary, we have demonstrated that combining all MRI features yielded highest age prediction accuracy in healthy individuals. Further, we found a varied prediction accuracy across the disease groups, suggesting metabolic and neurovascular aberrations in prodromal phases of neurodegenerative diseases and SZ. Together, these findings indicate that multidimensional neuroimaging of patients may provide a brain-based mapping of overlapping and distinct pathophysiology in common disorders of the brain using multimodal imaging.

## Data Availability

Due to ethical and data security issues related to the sensitive nature of the clinical data we are not allowed to share the data without specific IRB approval and data use agreements with the relevant institution. More information can be obtained through the corresponding author.

## Acknowledgements and funding

The study is supported by ERA-Net Cofund through the ERA PerMed project ‘IMPLEMENT’, the Research Council of Norway (223273, 249795, 248238, 276082, 286838, 298646, 300767), the South-Eastern Norway Regional Health Authority (2014097, 2015044, 2015073, 2016083, 2018037, 2018076, 2019101), the Norwegian ExtraFoundation for Health and Rehabilitation (2015/FO5146), the Novo Nordisk Foundation (NNF16OC0019856), KG Jebsen Stiftelsen, and the European Research Council under the European Union’s Horizon 2020 research and Innovation program (ERC StG, Grant 802998).

## Supplementary Material

### 1. Demographics

**Figure S1.**
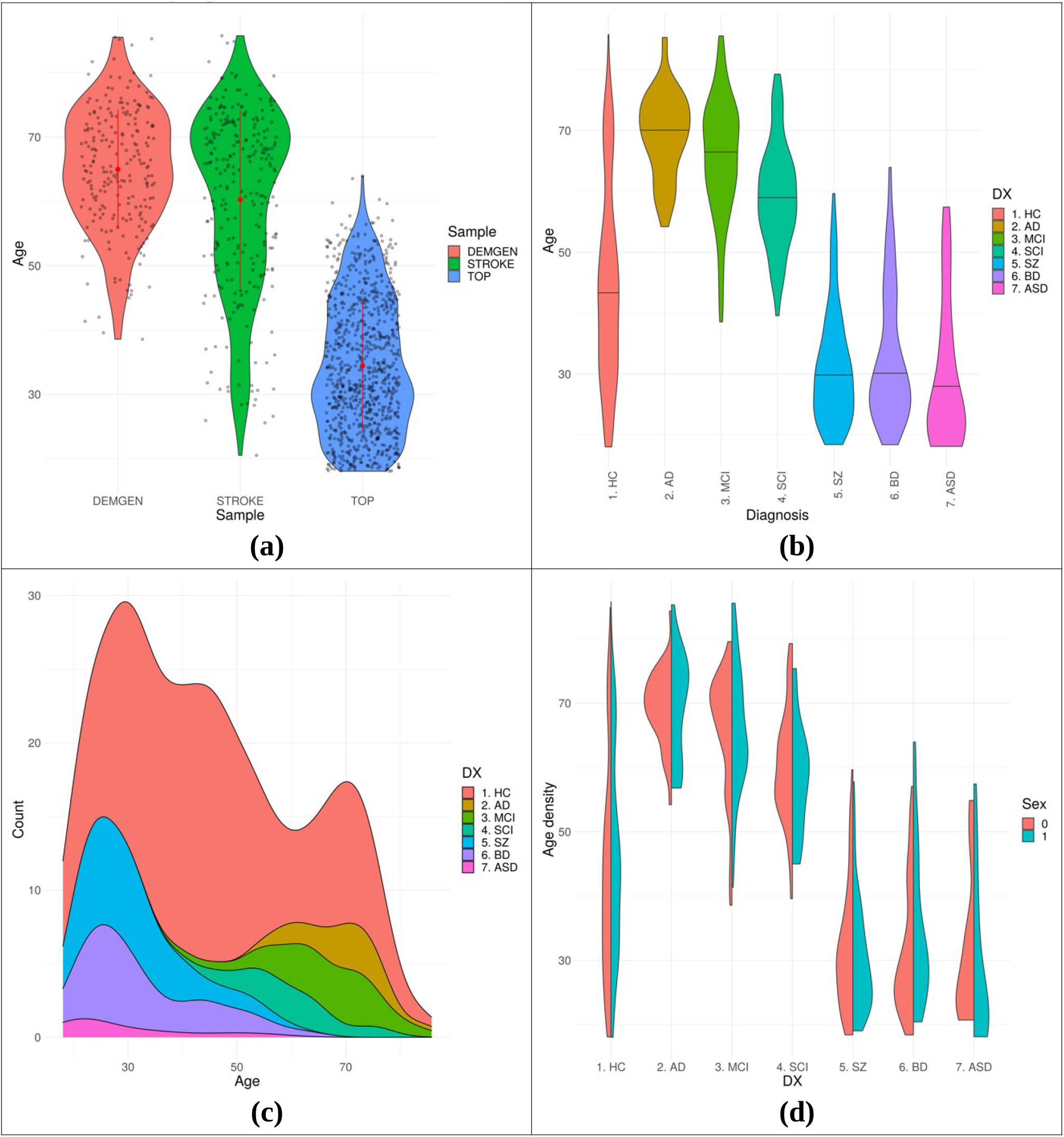
Data demographics separated by sample (a), and by diagnosis (b), cumulative distribution (c) and sex distribution for each diagnostic category (0-females, 1-males)

**Table S1.** Subjects demographics summarized by a sample

| Sample | n | Mean age | Sd age | % male | Min age | Max age |
| --- | --- | --- | --- | --- | --- | --- |
| DEMGEN | 197 | 64.9 | 9.5 | 47.2 | 38.6 | 85.5 |
| STROKE | 266 | 60.2 | 13.9 | 64.3 | 20.5 | 85.8 |
| TOP | 815 | 34.7 | 10.3 | 49.8 | 18.0 | 63.9 |

**Table S2.** Mean and standard deviation (SD) of clinical scores of participants with neurodevelopmental and neurodegenerative disorders. Sample size n is smaller, because scores were available only to subset of subjects with imaging data.

| Diagnosis | ASD | BD | SCZ | AD | MCI | SCI |
| --- | --- | --- | --- | --- | --- | --- |
| PANNS | Positive and Negative Syndrome Scale |  |  |  |  |  |
| N |  | 130 | 150 | NA | NA | NA |
| Mean |  | 45.0 | 59.0 | NA | NA | NA |
| SD |  | 9.5 | 15.3 | NA | NA | NA |
| GAF | Global Assessment of Functioning |  |  |  |  |  |
| N |  | 131 | 150 | NA | NA | NA |
| Mean |  | 25.6 | 30.8 | NA | NA | NA |
| SD |  | 5.4 | 7.5 | NA | NA | NA |
| IDS | Inventory of Depressive Symptoms |  |  |  |  |  |
| N |  | 129 | 113 | NA | NA | NA |
| Mean |  | 16.3 | 14.9 | NA | NA | NA |
| SD |  | 11.1 | 11.4 | NA | NA | NA |
| CDSS | Calgary Depression Scale for Schizophrenia |  |  |  |  |  |
| N |  | 127 | 139 | NA | NA | NA |
| Mean |  | 5.1 | 4.7 | NA | NA | NA |
| SD |  | 4.7 | 4.4 | NA | NA | NA |
| YMRS | Young Mania Rating Scale |  |  |  |  |  |
| N |  | 130 | 140 | NA | NA | NA |
| Mean |  | 2.5 | 3.7 | NA | NA | NA |
| SD |  | 3.6 | 4.5 | NA | NA | NA |
| MMSE | Mini-Mental State Examination |  |  |  |  |  |
| N |  | NA | NA | 24 | 25 | 22 |
| Mean |  | NA | NA | 23.1 | 27.5 | 29.1 |
| SD |  | NA | NA | 4.4 | 3.0 | 1.0 |

### 2. Brain aging by decade

**Figure S2.**
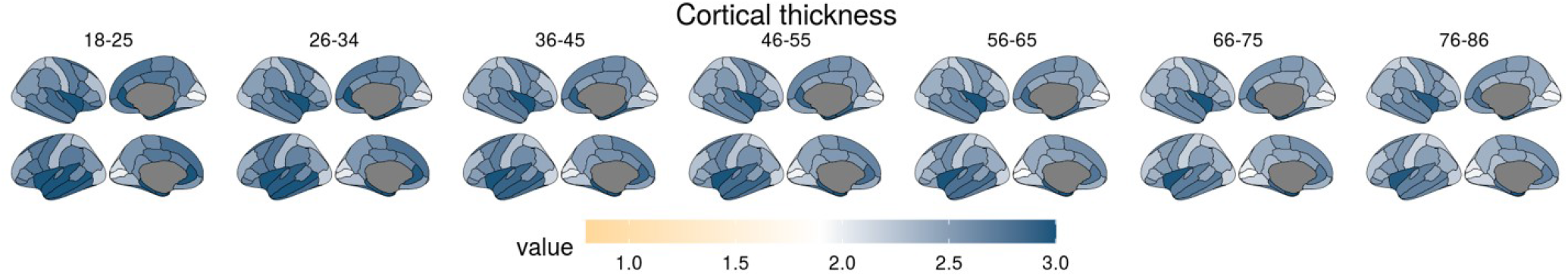
Mean cortical thickness changes (in mm) across lifespan summarized by decade

**Figure S3.**
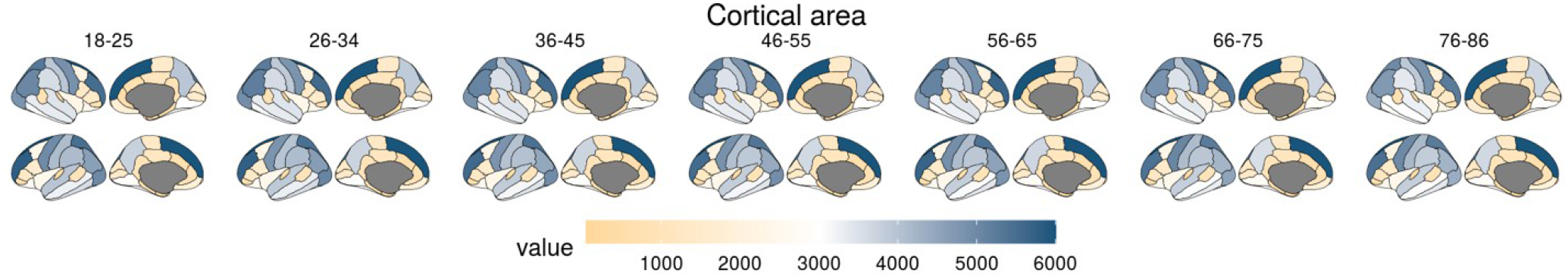
Mean cortical area changes (in mm2) across lifespan summarized by decade

**Figure S4.**
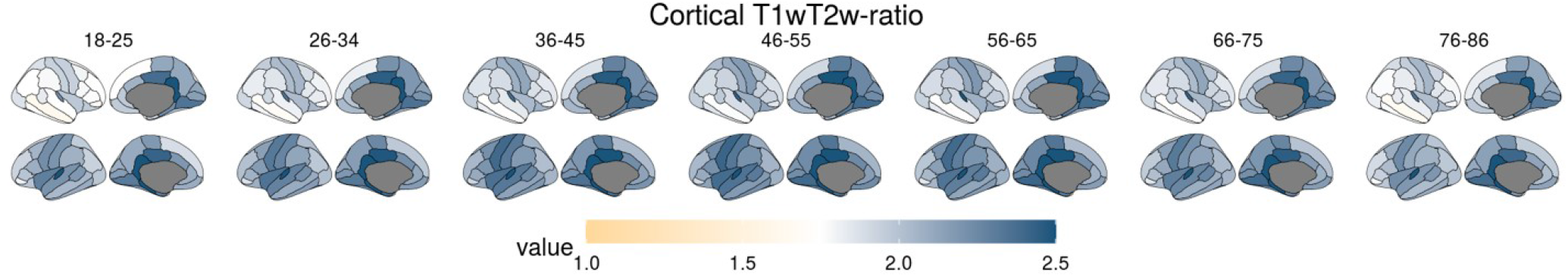
Mean cortical T1w/T2w ratio changes across lifespan summarized by decade

**Figure S5.**
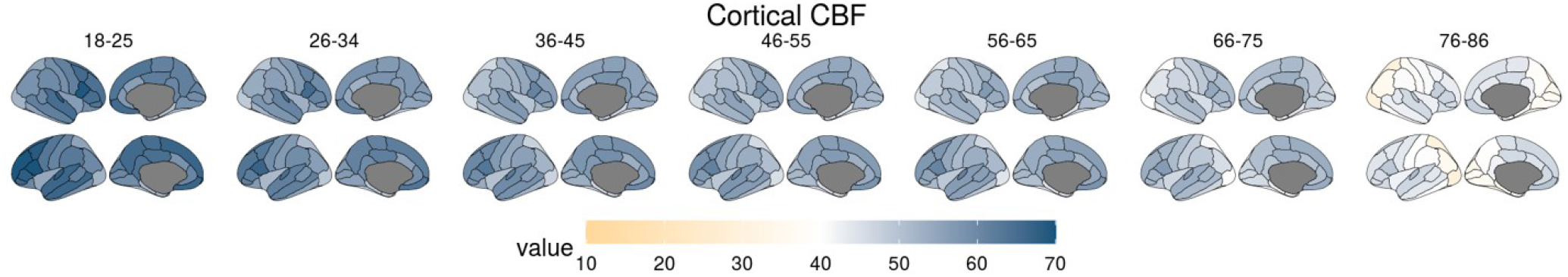
Mean cortical CBF changes (in ml/100g/min) across lifespan summarized by decade

**Figure S6.**
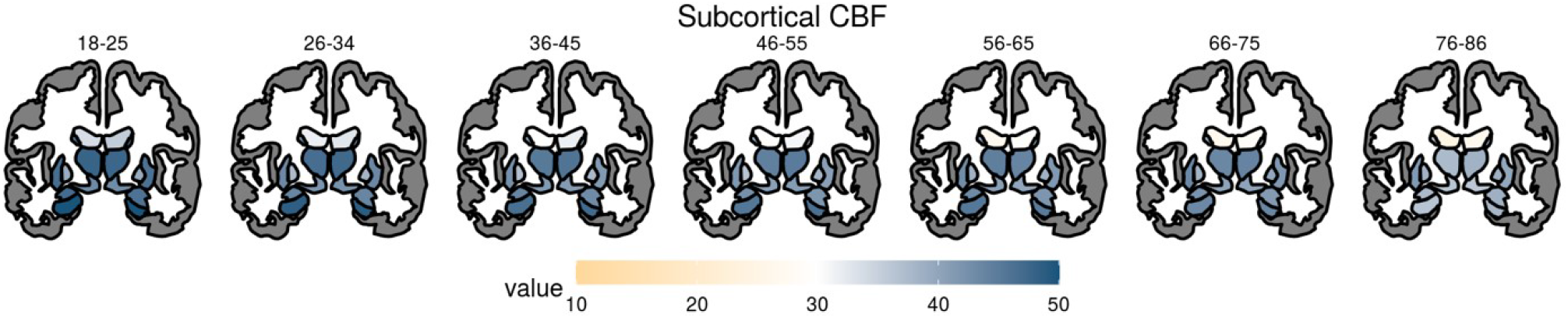
Mean subcortical CBF changes (in ml/100g/min) across lifespan summarized by decade

**Figure S7.**
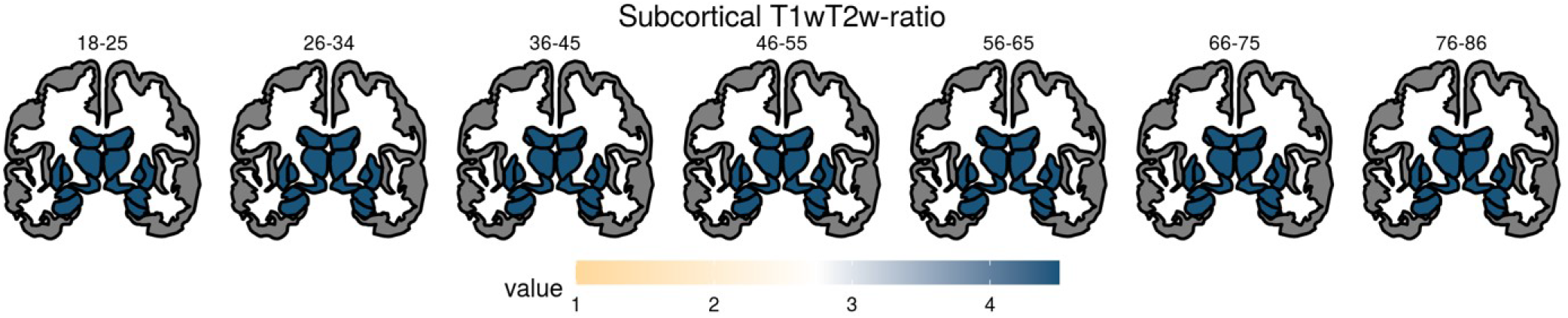
Mean subcortical T1w/T2w ratio changes across lifespan summarized by decade

**Figure S8.**
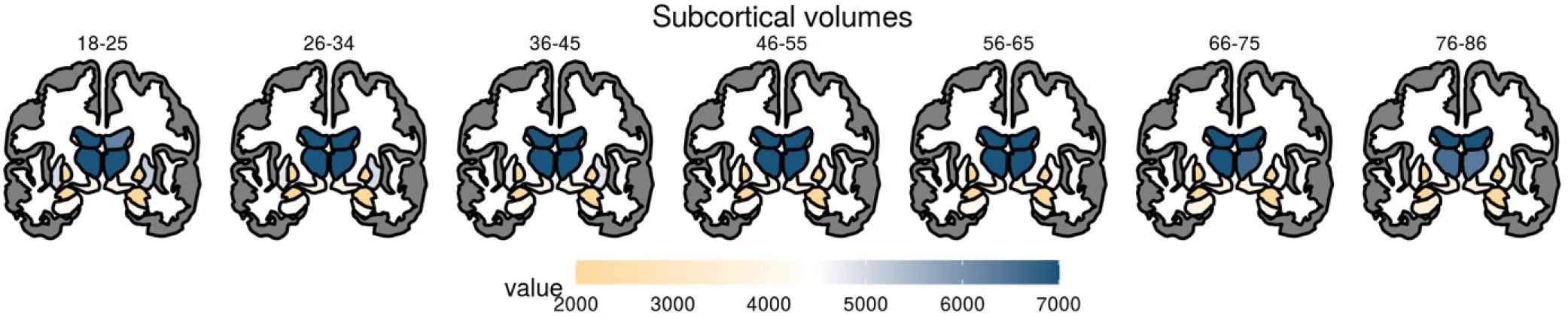
Mean subcortical volume changes (in mm3) across lifespan summarized by decade

### 3. Developmental trajectories

**Figure S9.**
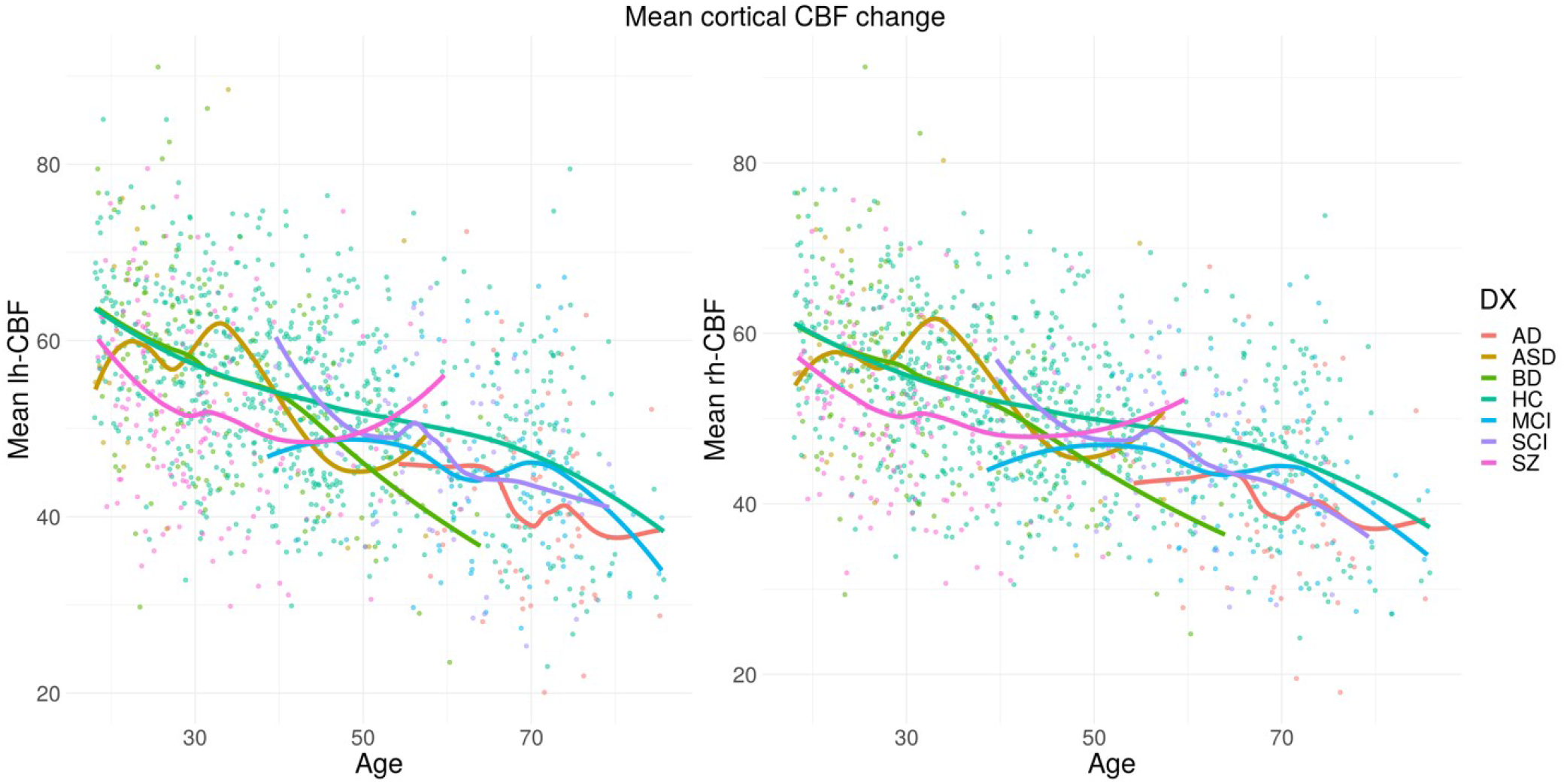
Developmental trajectories for mean cortical CBF (in ml/100g/min) for left and right hemisphere in. Trajectories for HC and patient groups were generated using loess function.

**Figure S10.**
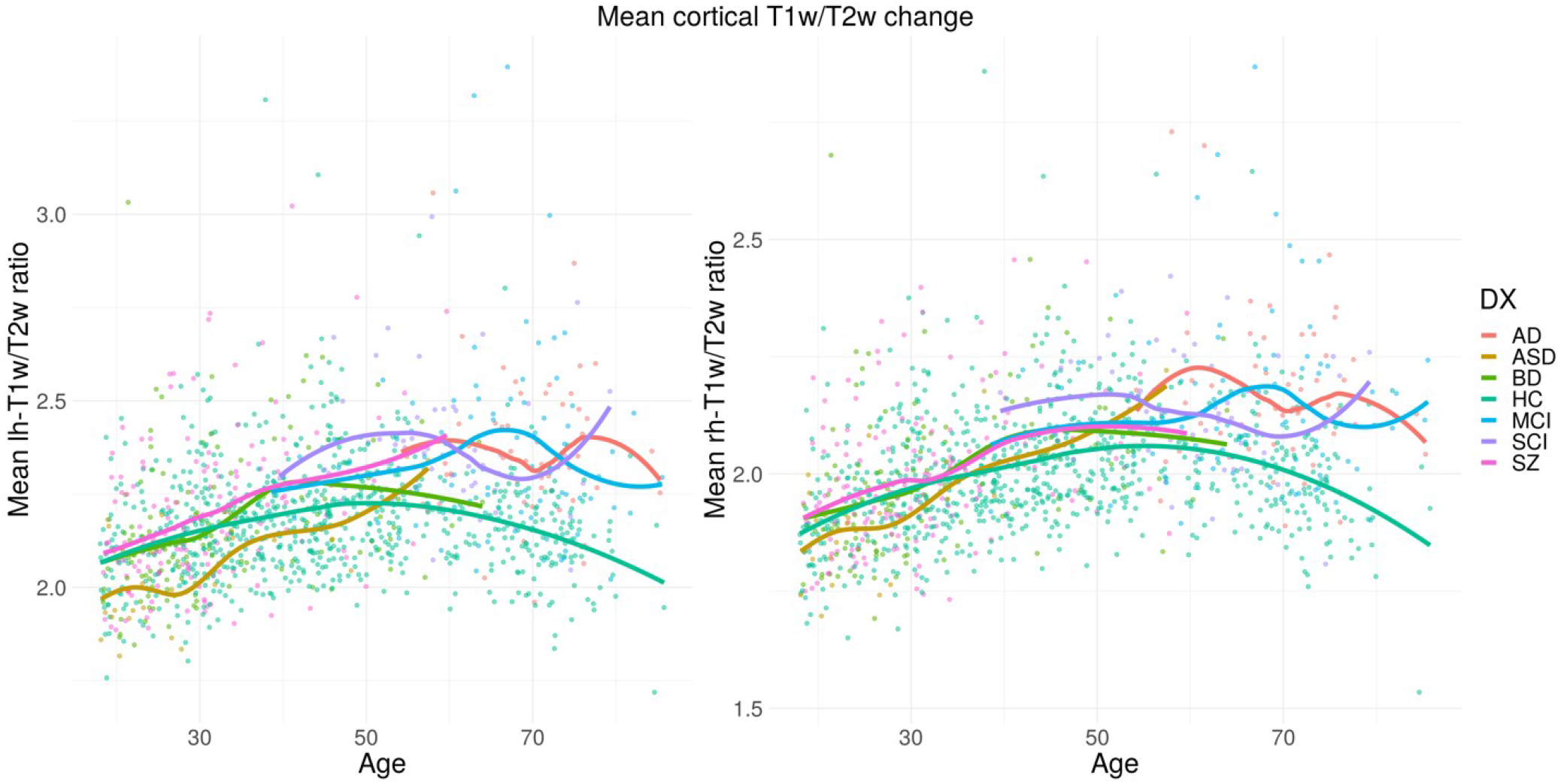
Developmental trajectories for mean cortical T1w/T2w ratio for left and right hemisphere. Trajectories for HC and patient groups were generated using loess function.

**Figure S11.**
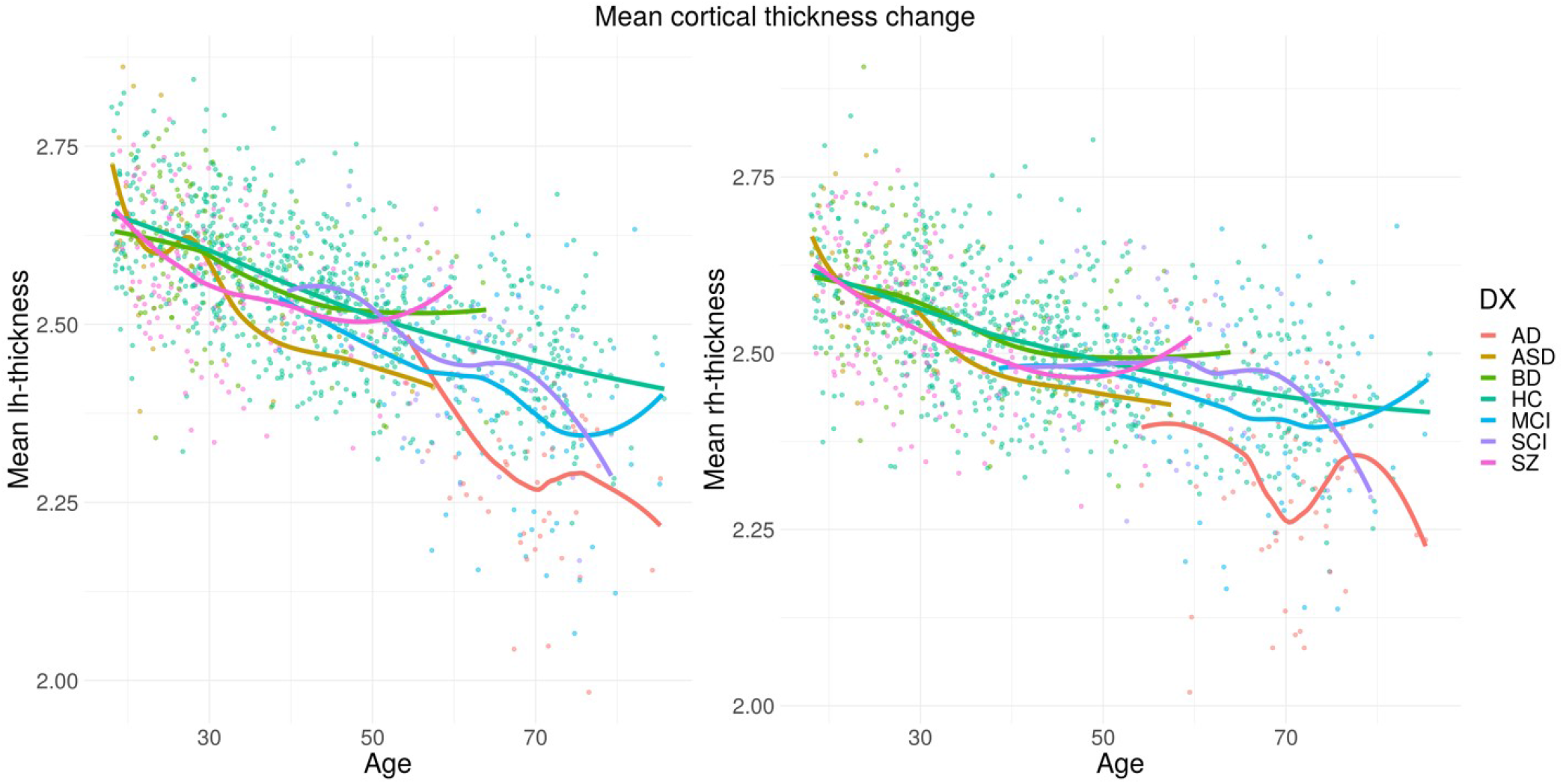
Developmental trajectories for mean cortical thickness (in mm) for left and right hemisphere. Trajectories for HC and patient groups were generated using loess function.

**Figure S12.**
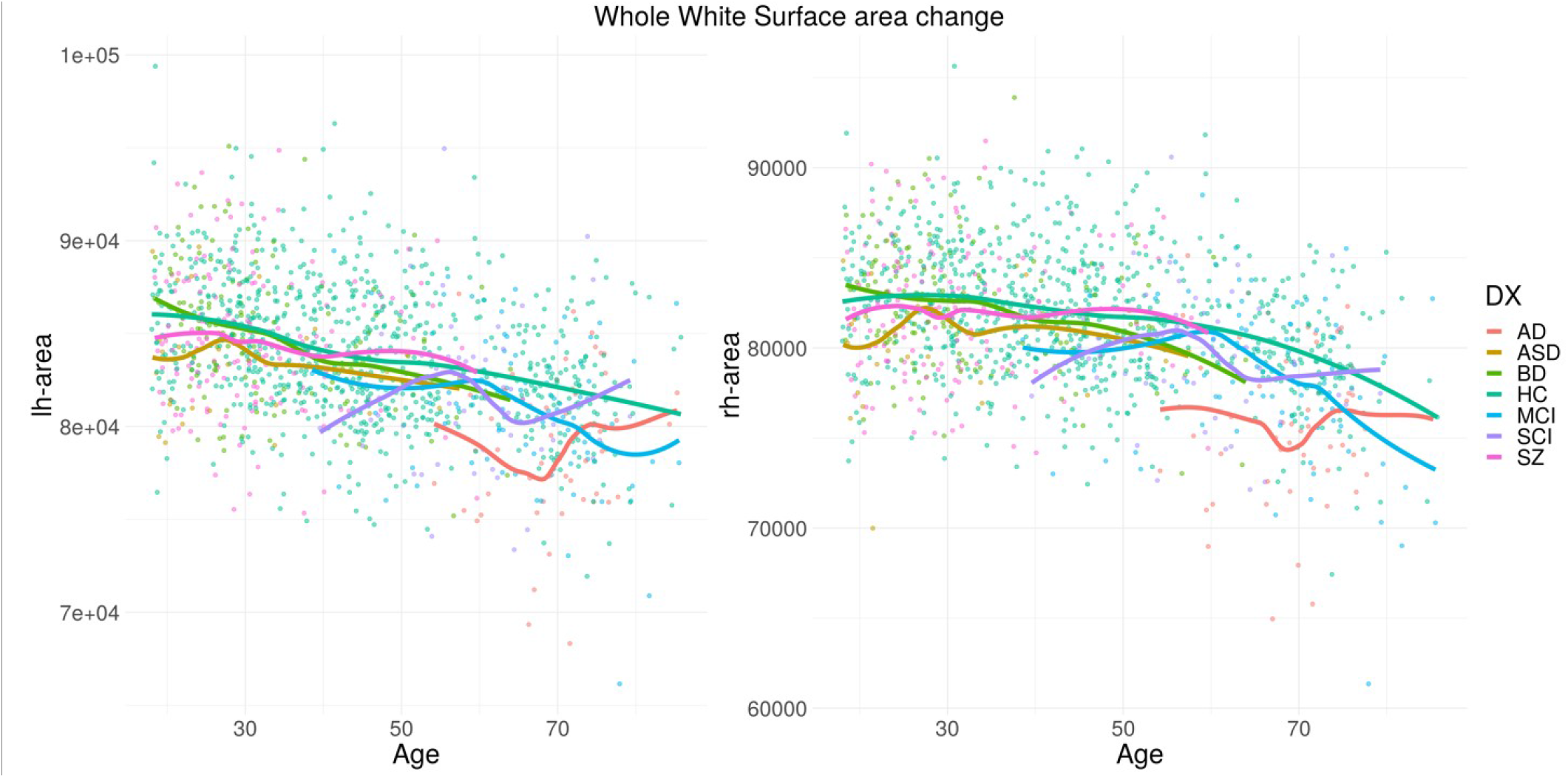
Developmental trajectories for whole white surface area (in mm2) for left and right hemisphere. Trajectories for HC and patient groups were generated using loess function.

**Figure S13.**
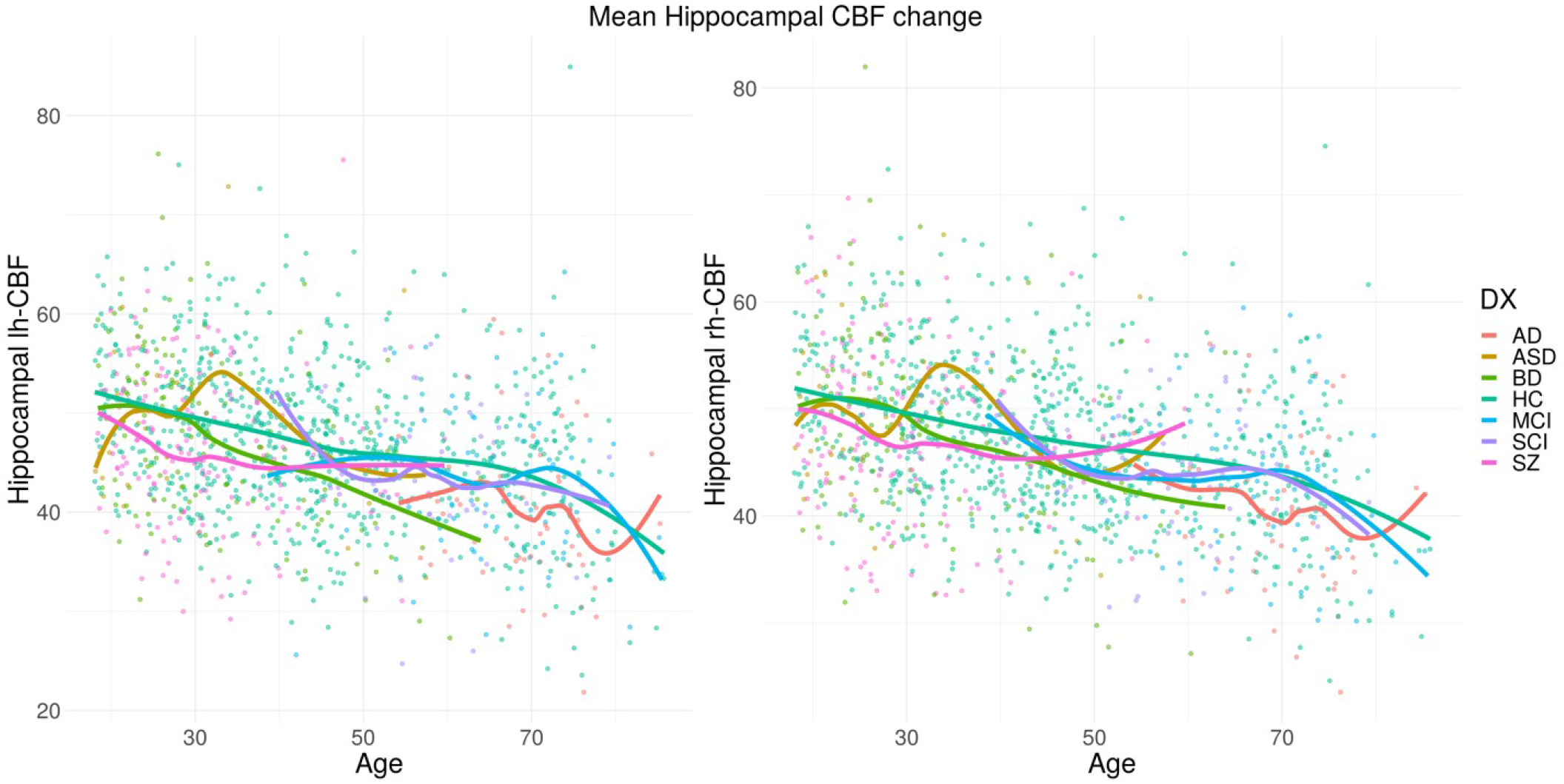
Developmental trajectories for mean hippocampal CBF (in ml/100g/min) for left and right hemisphere. Trajectories for HC and patient groups were generated using loess function.

**Figure S14.**
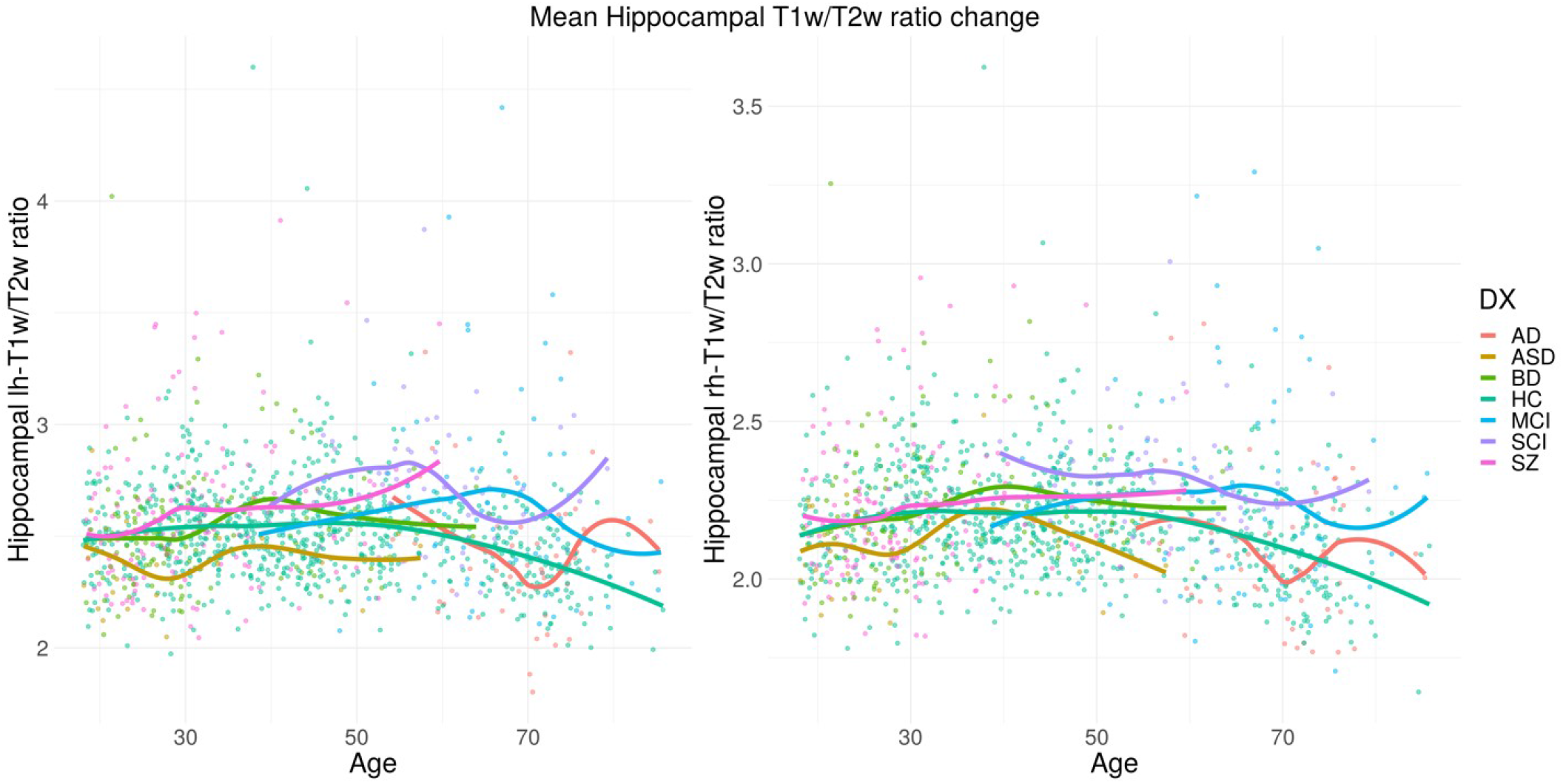
Developmental trajectories for mean hippocampal T1w/T2w ratio for left and right hemisphere. Trajectories for HC and patient groups were generated using loess function.

**Figure S15.**
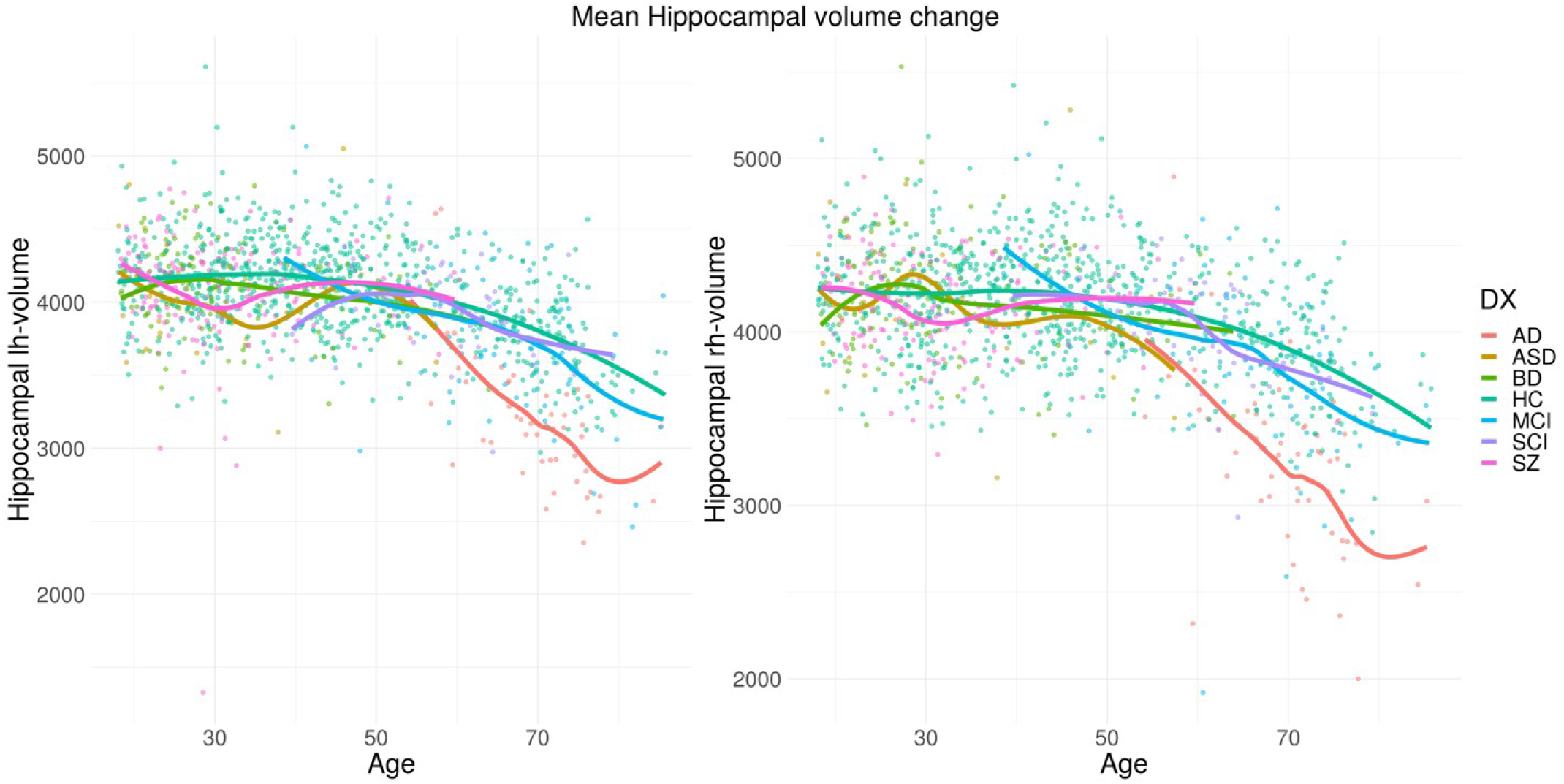
Developmental trajectories for mean hippocampal volume (in mm^3^) for left and right hemisphere. Trajectories for HC and patient groups were generated using loess function.

### 4. Brain structures used for brain age prediction

#### Cortical structures

1. Left bankssts 2. Left caudalanteriorcingulate 3. Left caudalmiddle-frontal 4. Left cuneus 5. Left entorhinal 6. Left fusiform 7. Left inferiorparietal 8. Left inferiortemporal 9. Left isthmuscingulate 10. Left lateraloccipital 11. Left lingual 12. Left lateralorbitofrontal 13. Left medialorbitofrontal 14. Left middletemporal 15. Left parahippocampal 16. Left paracentral 17. Left parsopercularis 18. Left parsorbitalis 19. Left parstriangularis 20. Left pericalcarine 21. Left postcentral 22. Left posteriorcingulate 23. Left precentral 24. Left precuneus 25. Left rostralanteriorcingulate 26. Left superiorfrontal 27. Left rostralmiddlefrontal 28. Left superiorparietal 29. Left superiortemporal 30. Left supramarginal 31. Left frontalpole 32. Left temporalpole 33. Left transversetemporal 34. Left in-sula 35. Right bankssts 36. Right caudalanteriorcingulate 37. Right caudalmiddlefrontal 38. Right cuneus 39. Right entorhinal 40. Right fusiform 41. Right inferiorparietal 42. Right inferiortemporal 43. Right isthmuscingulate 44. Right lateraloccipital 45. Right lateralorbitofrontal 46. Right lingual 47. Right medialorbitofrontal 48. Right middletemporal 49. Right parahippocampal 50. Right paracentral 51. Right parsopercularis 52. Right parsorbitalis 53. Right parstriangularis 54. Right pericalcarine 55. Right postcentral 56. Right posteriorcingulate 57. Right precentral 58. Right precuneus 59. Right rostralanteriorcingulate 60. Right rostralmiddlefrontal 61. Right superiorfrontal 62. Right superiorparietal 63. Right superiortemporal 64. Right supramarginal 65. Right frontalpole 66. Right temporalpole 67. Right transversetemporal 68. Right insula

#### Subcortical structures

1. Left Lateral Ventricle 2. Left Inf Lat Vent 3. Left Cerebellum White Matter 4. Left Cerebellum Cortex 5. Left Thalamus Proper 6. Left Caudate 7. Left Putamen 8. Left Pallidum 9. X3rd Ventricle 10. Brain Stem 11. Left Hippocampus 12. Left Amygdala 13. CSF 14. Left Accumbens area 15. Left VentralDC 16. Left choroid plexus 17. Right Lateral Ventricle 18. Right Inf Lat Vent 19. Right Cerebellum White Matter 20. Right Cerebellum Cortex 21. Right Thalamus Proper 22. Right Caudate 23. Right Putamen 24. Right Pallidum 25. Right Hippocampus 26. Right Amygdala 27. Right Accumbens area 28. Right VentralDC 29. Right choroid plexus 30. Optic Chiasm 31. CC Posterior 32. CC Mid Posterior 33. CC Central CC Mid Anterior 35. CC Anterior

### 5. Age Bias Compensation

**Figure S16.**
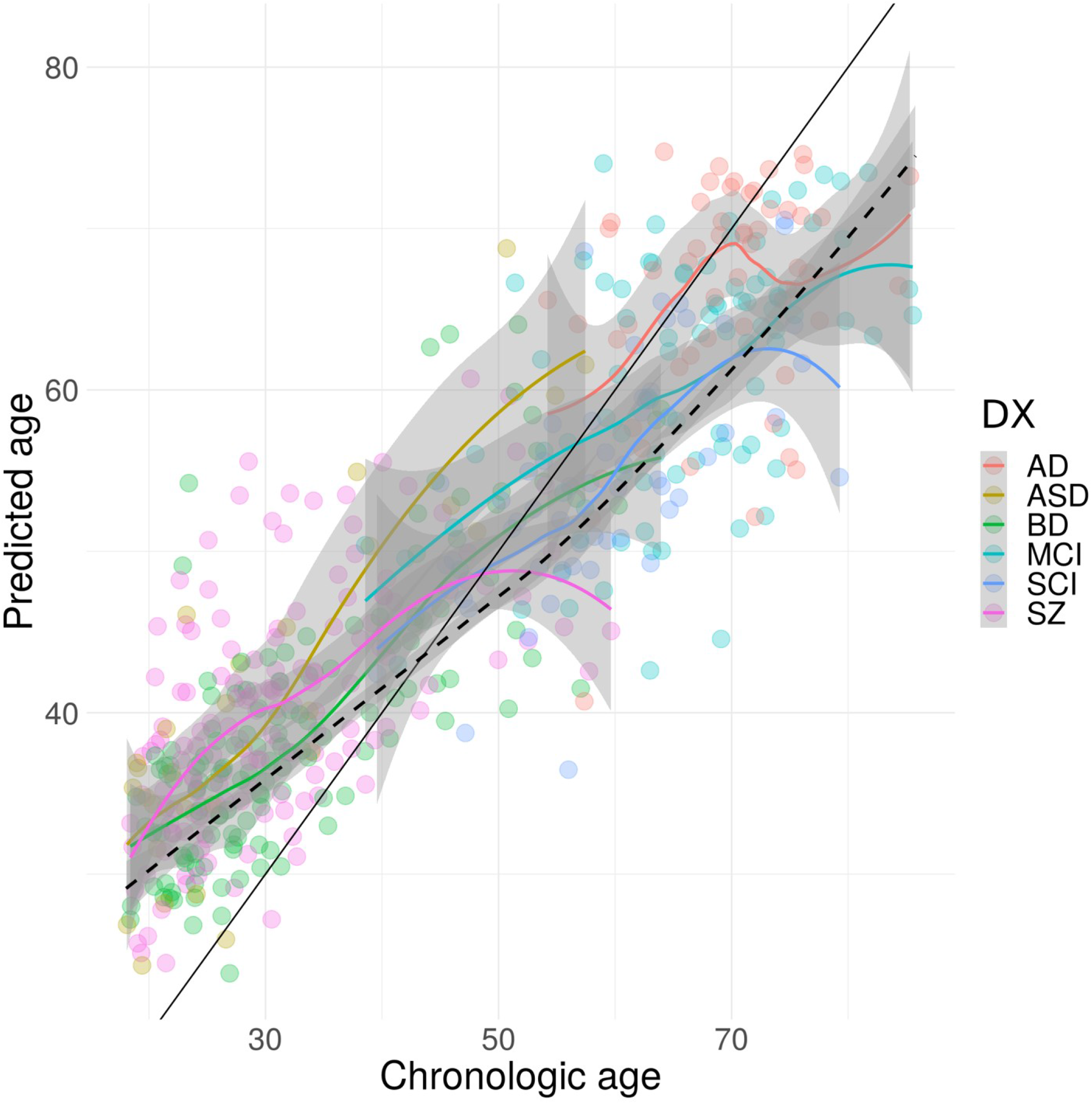
Dependency between chronological age vs machine predicted one based on all modalities. Color points and lines represent different patient groups. Dashed black line, represents HC, on whom model was trained.

**Figure S17.**
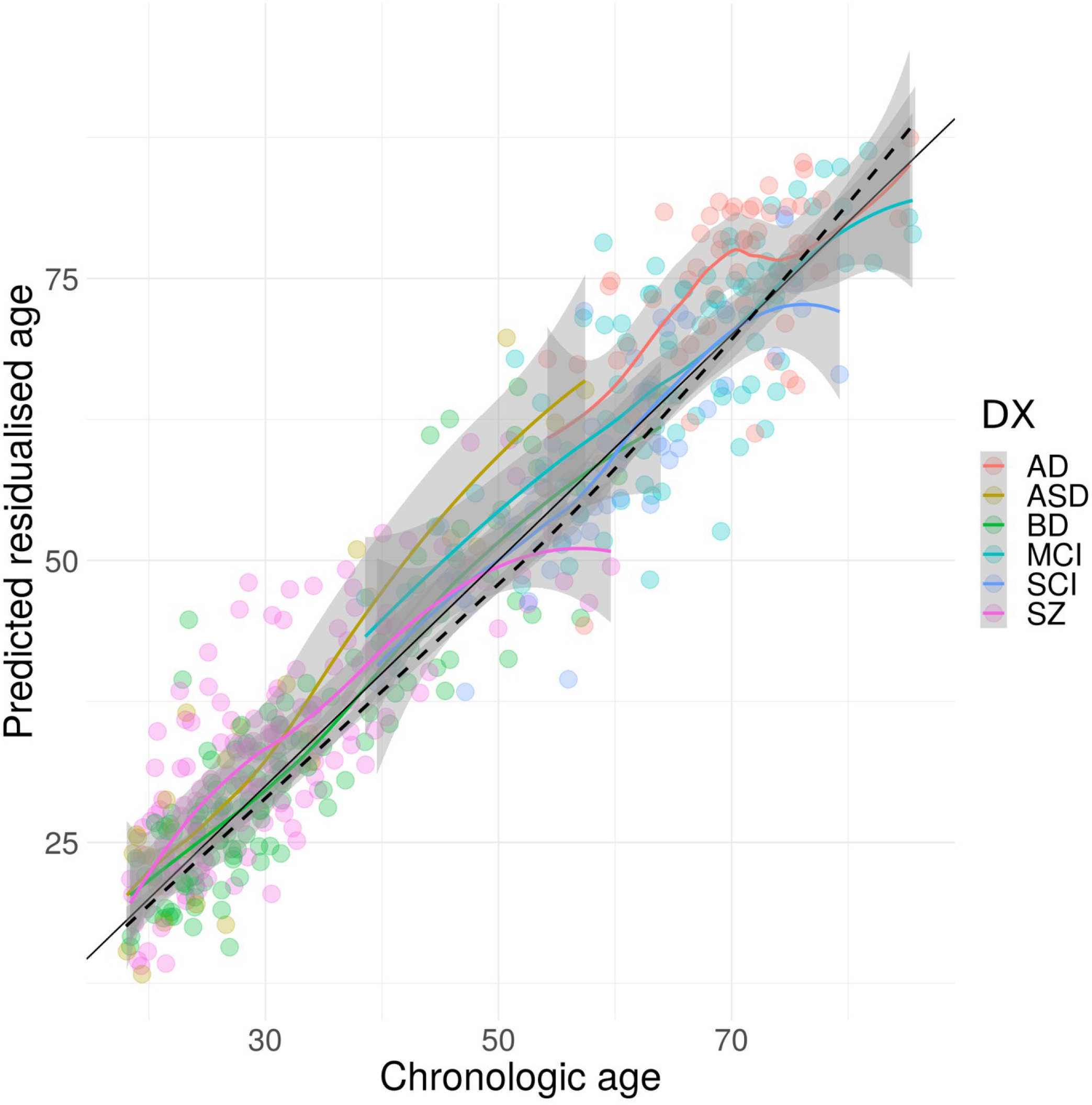
Dependency between chronological age vs machine predicted one with predicted age being residualized for age. Color points and lines represent different patient groups. Dashed black line, represents HC, on whom model was trained. Based on all modalities integrated.

### 6. Group Comparison Demographics

**Table S3.**
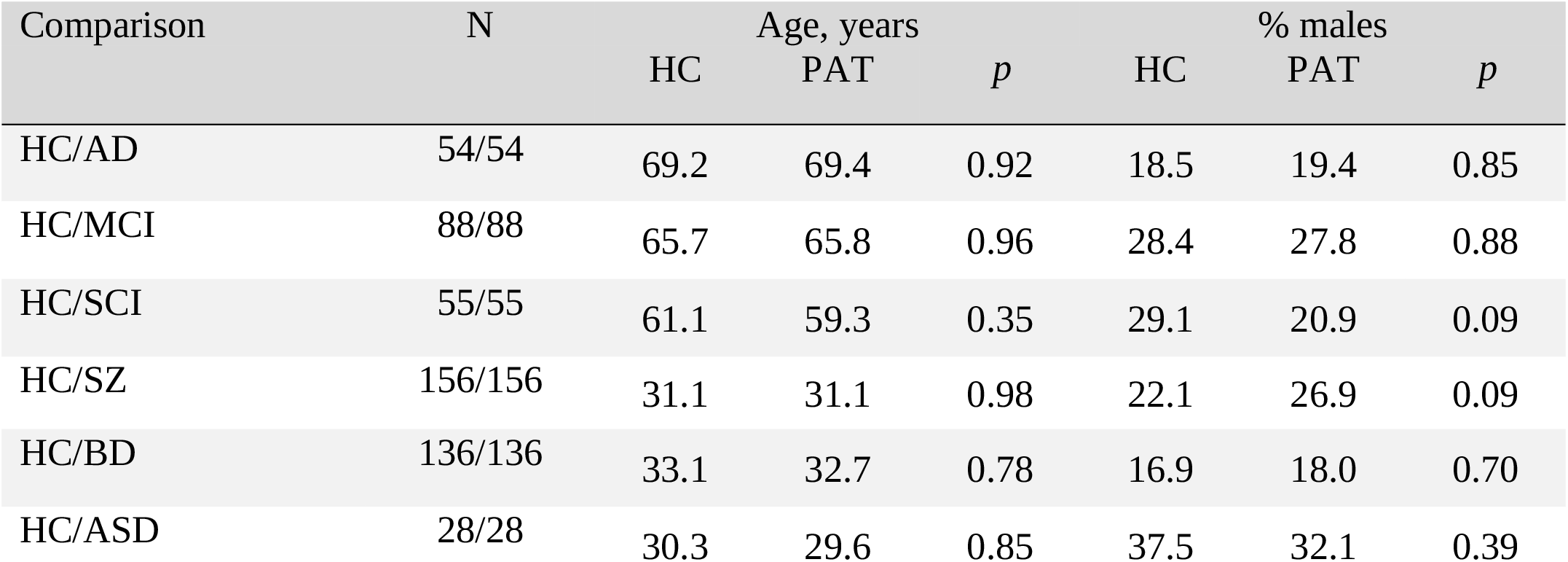
Subjects demographics for group comparison. Subjects were matched by the means of matchIt package using nearest neighbor matching with 1:1 ratio and logistic regression distance. Along with number of subjects in each group (N=54/54 meaning, that we have 54 HC and 54 AD subjects), mean age in each group, two group comparison p value and percentage of males within each group are shown. There are no significant differences neither in age nor in sex in any of the comparisons

### 7. Model fit in healthy controls

**Table S4.** Fit in health control population with r^2^ of a model fit, mean square error (mae) and root mean square error (rmse) given for each modality.

| Modality | $r^2$ | mae | rmse |
| --- | --- | --- | --- |
| Global multimodal | 0.77 | 6.5 | 8.2 |
| Global T1w/T2w-ratio | 0.73 | 6.9 | 8.7 |
| Global T1w-based | 0.72 | 6.9 | 8.8 |
| Subcortical T1w/T2w-ratio | 0.68 | 7.4 | 9.4 |
| Subcortical volumes | 0.67 | 7.5 | 9.4 |
| Cortical thickness | 0.58 | 8.4 | 10.5 |
| Global CBF | 0.51 | 9.2 | 11.5 |
| Subcortical CBF | 0.48 | 9.4 | 11.8 |
| Cortical T1w/T2w-ratio | 0.39 | 10.2 | 12.9 |
| Cortical area | 0.31 | 11.2 | 13.6 |
| Cortical CBF | 0.31 | 10.8 | 13.4 |

### 8. Feature Importance

**Figure S18.**
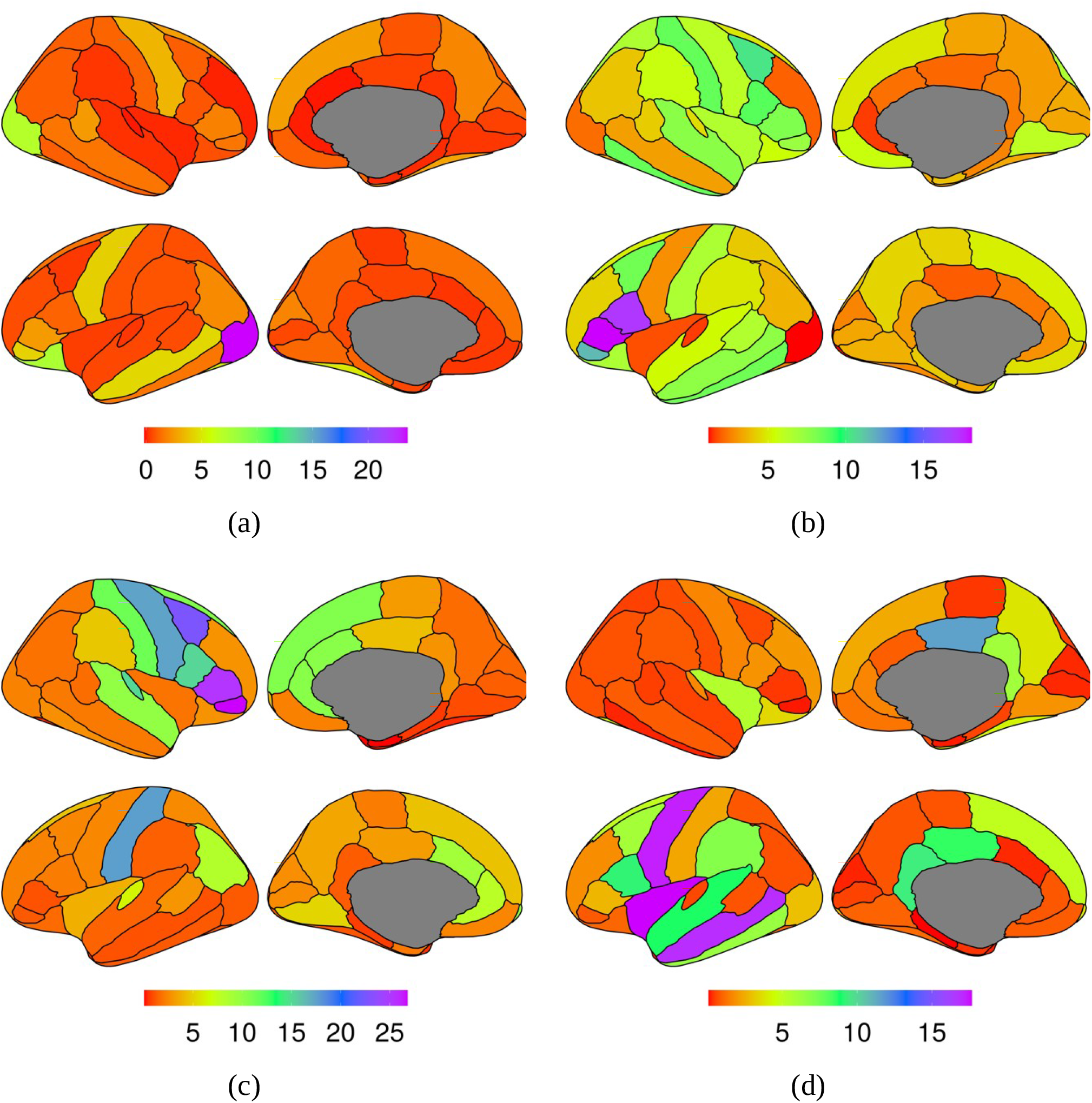
Feature importance measured as increase in MSE (left) shown as a colormap overlaid on the brain for cortical area (a) CBF (b) T1w/T2w ratio (c) and thickness (d)

**Figure S19.**
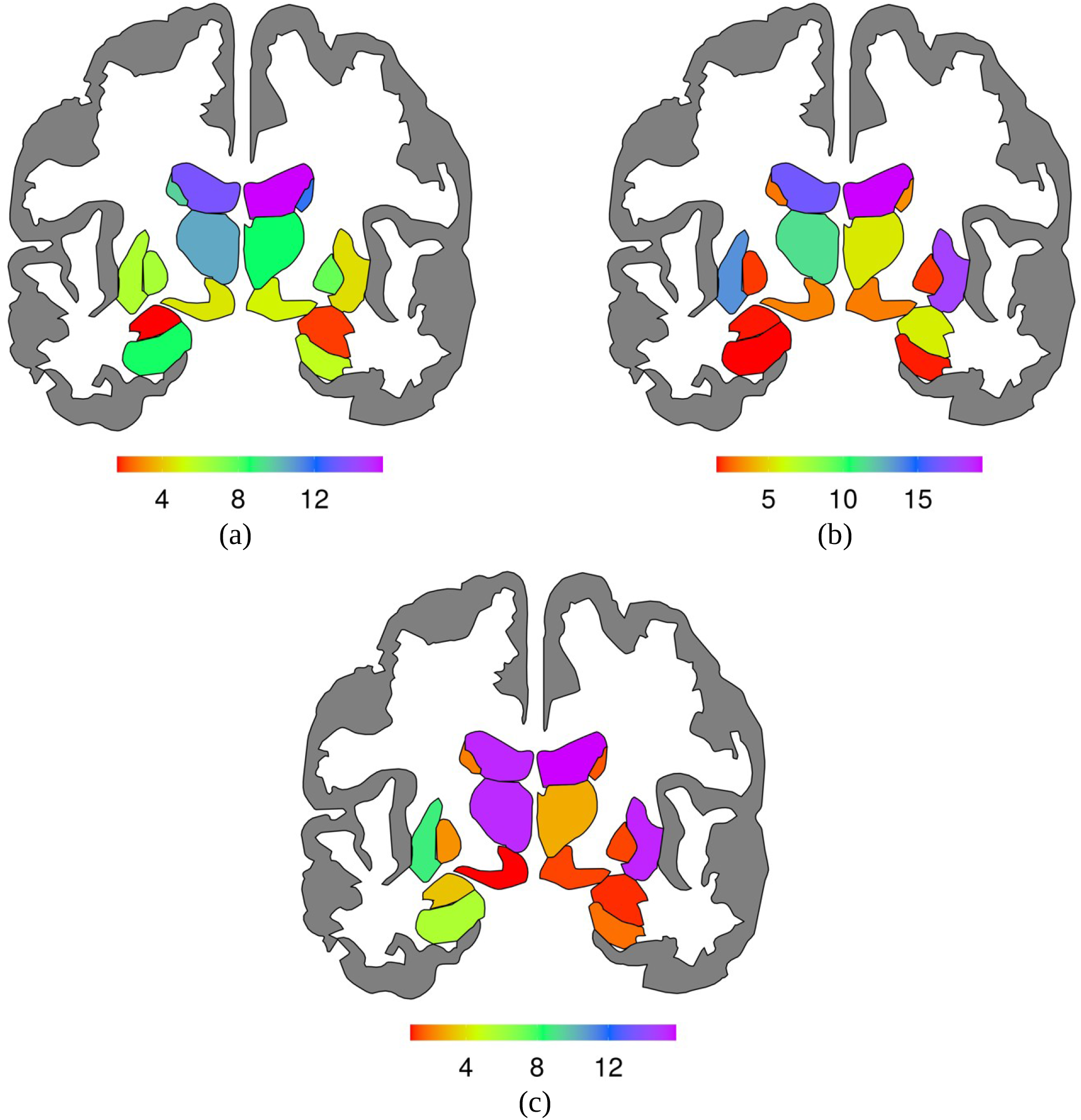
Feature importance measured as increase in MSE (left) shown as a colormap overlaid on the brain for subcortical CBF (a) T1w/T2w ratio (b) and volumes (c)

### 9. Feature importance

**Table S5.** Feature importance for the model (everything) with best age fir r2=0.77 in HC. Only top 20 most informative features are shown.

| Strucutre | %IncMSE | IncNodePurity | Part | Mod | Hemisphere |
| --- | --- | --- | --- | --- | --- |
| X3rd Ventricle | 29.5 | 19,762.6 | sub | T1w/T2w |  |
| X3rd Ventricle | 22.7 | 16,573.4 | sub | vol |  |
| Insula | 8.6 | 7,994.7 | cort | thickness | lh |
| Lateral Ventricle | 6.0 | 5,218.1 | sub | vol | lh |
| Thalamus Proper | 5.8 | 6,121.5 | sub | vol | lh |
| Lateral Ventricle | 5.7 | 6,093.8 | sub | vol | rh |
| CC Anterior | 5.6 | 3,872.7 | sub | T1w/T2w |  |
| Accumbens | 5.1 | 6,340.5 | sub | vol | rh |
| Middletemporal | 4.8 | 4,964.7 | cort | thickness | lh |
| Choroid Plexus | 4.6 | 4,422.9 | sub | vol | lh |
| Precentral | 4.1 | 4,195.4 | cort | thickness | lh |
| Parsopercularis | 3.8 | 3,104.3 | cort | thickness | lh |
| Inferiortemporal | 3.8 | 1,776.6 | cort | thickness | lh |
| CSF | 3.2 | 2,697.8 | sub | T1w/T2w |  |
| Accumbens | 3.0 | 2,312.0 | sub | vol | lh |
| Putamen | 2.9 | 2,359.5 | sub | vol | rh |
| Posteriorcingulate | 2.6 | 2,807.8 | cort | thickness | rh |
| Superiortemporal | 2.6 | 2,408.1 | cort | thickness | lh |
| Precentral | 2.5 | 1,694.1 | cort | T1w/T2w | rh |

### 10. BAG

**Table S6.** Mean group differences between patients and age and sex matched HC. BAG was additionally controlled for sex and age within each comparison, using linear models and p values were adjusted for 66 comparisons using FDR. Cd stands for Cohen’s d.

| Modality | BAG<br>diff | Cd | p_fdr |  | BAG diff | Cd | p_fdr |  |
| --- | --- | --- | --- | --- | --- | --- | --- | --- |
| **AD** |  |  |  |  | **SZ** |  |  |  |
| Cortical area | 6.2 | 0.98 | 1.33E-05 | *** | 3.1 | 0.56 | 1.25E-05 | *** |
| Cortical CBF | 6.8 | 0.76 | 6.87E-04 | *** | 4.2 | 0.61 | 2.80E-06 | *** |
| Cortical T1w/T2w-ratio | 0.5 | 0.10 | 6.89E-01 |  | 0.8 | 0.16 | 2.23E-01 |  |
| Cortical thickness | 6.9 | 0.96 | 2.11E-05 | *** | 3.2 | 0.49 | 1.03E-04 | *** |
| Global CBF | 6.4 | 0.77 | 5.94E-04 | *** | 4.5 | 0.68 | 3.57E-07 | *** |
| Global multimodal | 5.8 | 0.86 | 1.28E-04 | *** | 3.5 | 0.65 | 6.97E-07 | *** |
| Global T1w-based | 7.4 | 1.09 | 2.94E-06 | *** | 3.0 | 0.56 | 1.25E-05 | *** |
| Global T1w/T2w-ratio | 2.9 | 0.47 | 3.28E-02 | * | 1.7 | 0.30 | 1.63E-02 | * |
| Subcortical CBF | 4.5 | 0.52 | 1.87E-02 | * | 3.5 | 0.55 | 1.33E-05 | *** |
| Subcortical T1w/T2w-ratio | 3.1 | 0.41 | 6.06E-02 |  | 1.7 | 0.29 | 1.94E-02 | * |
| Subcortical volumes | 7.9 | 1.04 | 7.17E-06 | *** | 0.8 | 0.14 | 2.67E-01 |  |
| **MCI** |  |  |  |  | **BD** |  |  |  |
| Cortical area | 3.0 | 0.47 | 6.53E-03 | ** | 2.6 | 0.44 | 1.20E-03 | ** |
| Cortical CBF | 4.0 | 0.45 | 9.19E-03 | ** | 2.4 | 0.36 | 8.36E-03 | ** |
| Cortical T1w/T2w-ratio | -0.9 | 0.17 | 3.15E-01 |  | 1.1 | 0.21 | 1.21E-01 |  |
| Cortical thickness | 1.2 | 0.17 | 3.15E-01 |  | 1.8 | 0.30 | 2.59E-02 | * |
| Global CBF | 3.7 | 0.47 | 6.53E-03 | ** | 2.8 | 0.42 | 1.78E-03 | ** |
| Global multimodal | 1.8 | 0.29 | 9.08E-02 |  | 2.2 | 0.43 | 1.64E-03 | ** |
| Global T1w-based | 2.4 | 0.34 | 4.30E-02 | * | 2.5 | 0.48 | 4.87E-04 | *** |
| Global T1w/T2w-ratio | 0.5 | 0.07 | 7.12E-01 |  | 1.0 | 0.17 | 2.14E-01 |  |
| Subcortical CBF | 3.2 | 0.42 | 1.47E-02 | * | 3.0 | 0.48 | 4.44E-04 | *** |
| Subcortical T1w/T2w-ratio | 0.3 | 0.03 | 8.53E-01 |  | 1.2 | 0.20 | 1.32E-01 |  |
| Subcortical volumes | 2.7 | 0.34 | 4.30E-02 | * | 1.4 | 0.24 | 7.72E-02 |  |
| **SCI** |  |  |  |  | **ASD** |  |  |  |
| Cortical area | -0.7 | 0.10 | 6.68E-01 |  | 2.1 | 0.31 | 3.15E-01 |  |
| Cortical CBF | 1.7 | 0.21 | 3.15E-01 |  | 2.2 | 0.36 | 2.43E-01 |  |
| Cortical T1w/T2w-ratio | -1.9 | 0.39 | 6.85E-02 |  | 2.8 | 0.51 | 9.68E-02 |  |
| Cortical thickness | 0.0 | 0.01 | 9.74E-01 |  | 6.5 | 1.00 | 1.64E-03 | ** |
| Global CBF | 3.6 | 0.47 | 2.71E-02 | * | 2.5 | 0.47 | 1.22E-01 |  |
| Global multimodal | 0.7 | 0.13 | 5.88E-01 |  | 4.9 | 0.97 | 1.97E-03 | ** |
| Global T1w-based | 0.5 | 0.08 | 7.22E-01 |  | 6.7 | 1.19 | 2.56E-04 | *** |
| Global T1w/T2w-ratio | 0.2 | 0.04 | 8.53E-01 |  | 3.5 | 0.63 | 4.18E-02 | * |
| Subcortical CBF | 4.1 | 0.54 | 1.23E-02 | * | 4.2 | 0.77 | 1.41E-02 | * |
| Subcortical T1w/T2w-ratio | 0.1 | 0.02 | 9.40E-01 |  | 4.0 | 0.70 | 2.35E-02 | * |
| Subcortical volumes | 0.6 | 0.08 | 7.12E-01 |  | 5.8 | 0.99 | 1.78E-03 | ** |

**Figure S20.**
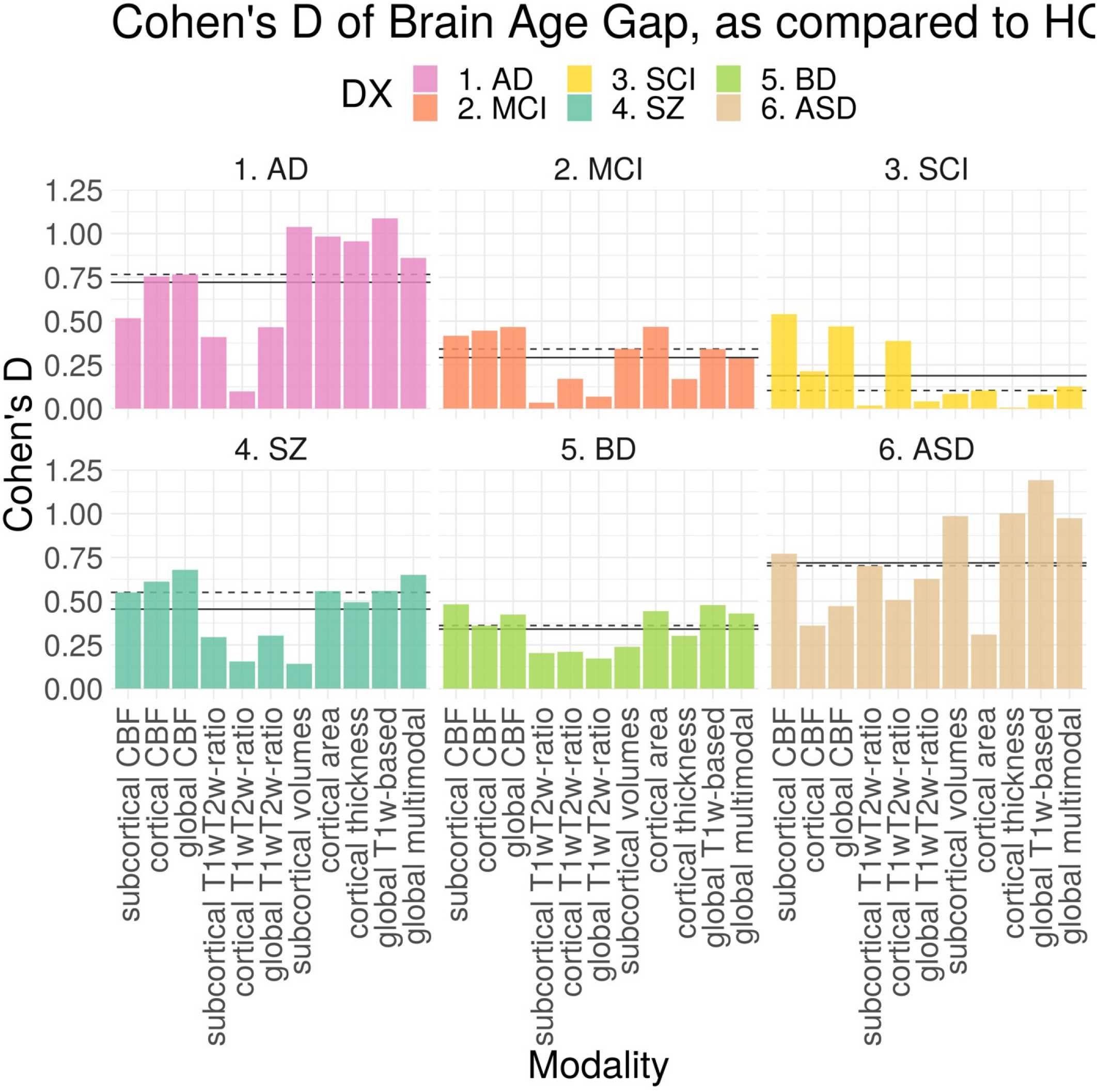
Effect sizes for each modality and diagnosis. Mean (solid) and median(dashed) per diagnosis are marked by lines.

### 11. AUC, sensitivity and specificity

**Table S7.** AUC estimated each modality and patient group.

|  | sub-CBF | cortical CBF | global CBF | sub-T1w/T2w-ratio | cortical T1w/T2w-ratio | global T1w/T2w-ratio | sub-volume | cortical thickness | cortical area | global T1w-based | global multimodal |
| --- | --- | --- | --- | --- | --- | --- | --- | --- | --- | --- | --- |
| AD | 0.64 | 0.71 | 0.72 | 0.60 | 0.52 | 0.64 | 0.77 | 0.75 | 0.76 | 0.81 | 0.74 |
| MCI | 0.61 | 0.64 | 0.63 | 0.52 | 0.56 | 0.53 | 0.59 | 0.54 | 0.64 | 0.60 | 0.57 |
| SCI | 0.65 | 0.60 | 0.63 | 0.51 | 0.62 | 0.51 | 0.52 | 0.51 | 0.54 | 0.52 | 0.52 |
| SZ | 0.64 | 0.66 | 0.68 | 0.59 | 0.56 | 0.59 | 0.53 | 0.64 | 0.63 | 0.65 | 0.68 |
| BD | 0.63 | 0.59 | 0.61 | 0.57 | 0.57 | 0.56 | 0.56 | 0.58 | 0.60 | 0.63 | 0.63 |
| ASL | 0.70 | 0.60 | 0.62 | 0.69 | 0.66 | 0.67 | 0.74 | 0.75 | 0.58 | 0.80 | 0.78 |

**Table S8.** Sensitivity estimated each modality and patient group.

|  | sub-CBF | cortical CBF | global CBF | sub-T1w/T2w-ratio | cortical T1w/T2w-ratio | global T1w/T2w-ratio | sub-volume | cortical thickness | cortical area | global T1w-based | global multimodal |
| --- | --- | --- | --- | --- | --- | --- | --- | --- | --- | --- | --- |
| AD | 0.76 | 0.83 | 0.57 | 0.59 | 0.41 | 0.70 | 0.89 | 0.57 | 0.69 | 0.78 | 0.54 |
| MCI | 0.69 | 0.70 | 0.59 | 0.40 | 0.95 | 0.59 | 0.56 | 0.85 | 0.70 | 0.67 | 0.39 |
| SCI | 0.67 | 0.76 | 0.69 | 0.85 | 0.11 | 0.25 | 0.64 | 0.78 | 0.13 | 0.96 | 0.96 |
| SZ | 0.65 | 0.49 | 0.60 | 0.80 | 0.76 | 0.49 | 0.34 | 0.83 | 0.47 | 0.57 | 0.60 |
| BD | 0.66 | 0.43 | 0.71 | 0.78 | 0.57 | 0.68 | 0.56 | 0.62 | 0.35 | 0.76 | 0.56 |
| ASL | 0.86 | 0.57 | 0.54 | 0.71 | 0.96 | 0.68 | 0.82 | 0.68 | 0.43 | 0.82 | 0.71 |

**Table S9.** Specificity estimated each modality and patient group

|  | sub-CBF | cortical CBF | global CBF | sub-T1w/T2w-ratio | cortical T1w/T2w-ratio | global T1w/T2w-ratio | sub-volume | cortical thickness | cortical area | global T1w-based | global multimodal |
| --- | --- | --- | --- | --- | --- | --- | --- | --- | --- | --- | --- |
| AD | 0.52 | 0.57 | 0.80 | 0.59 | 0.70 | 0.63 | 0.54 | 0.83 | 0.85 | 0.72 | 0.87 |
| MCI | 0.51 | 0.53 | 0.66 | 0.73 | 0.10 | 0.57 | 0.60 | 0.28 | 0.55 | 0.52 | 0.77 |
| SCI | 0.58 | 0.53 | 0.55 | 0.24 | 0.91 | 0.84 | 0.47 | 0.36 | 0.96 | 0.18 | 0.20 |
| SZ | 0.58 | 0.78 | 0.70 | 0.38 | 0.35 | 0.69 | 0.78 | 0.38 | 0.81 | 0.70 | 0.70 |
| BD | 0.60 | 0.73 | 0.50 | 0.36 | 0.60 | 0.45 | 0.57 | 0.56 | 0.86 | 0.49 | 0.69 |
| ASL | 0.50 | 0.64 | 0.75 | 0.71 | 0.39 | 0.71 | 0.61 | 0.86 | 0.82 | 0.79 | 0.86 |

**Table S9.**
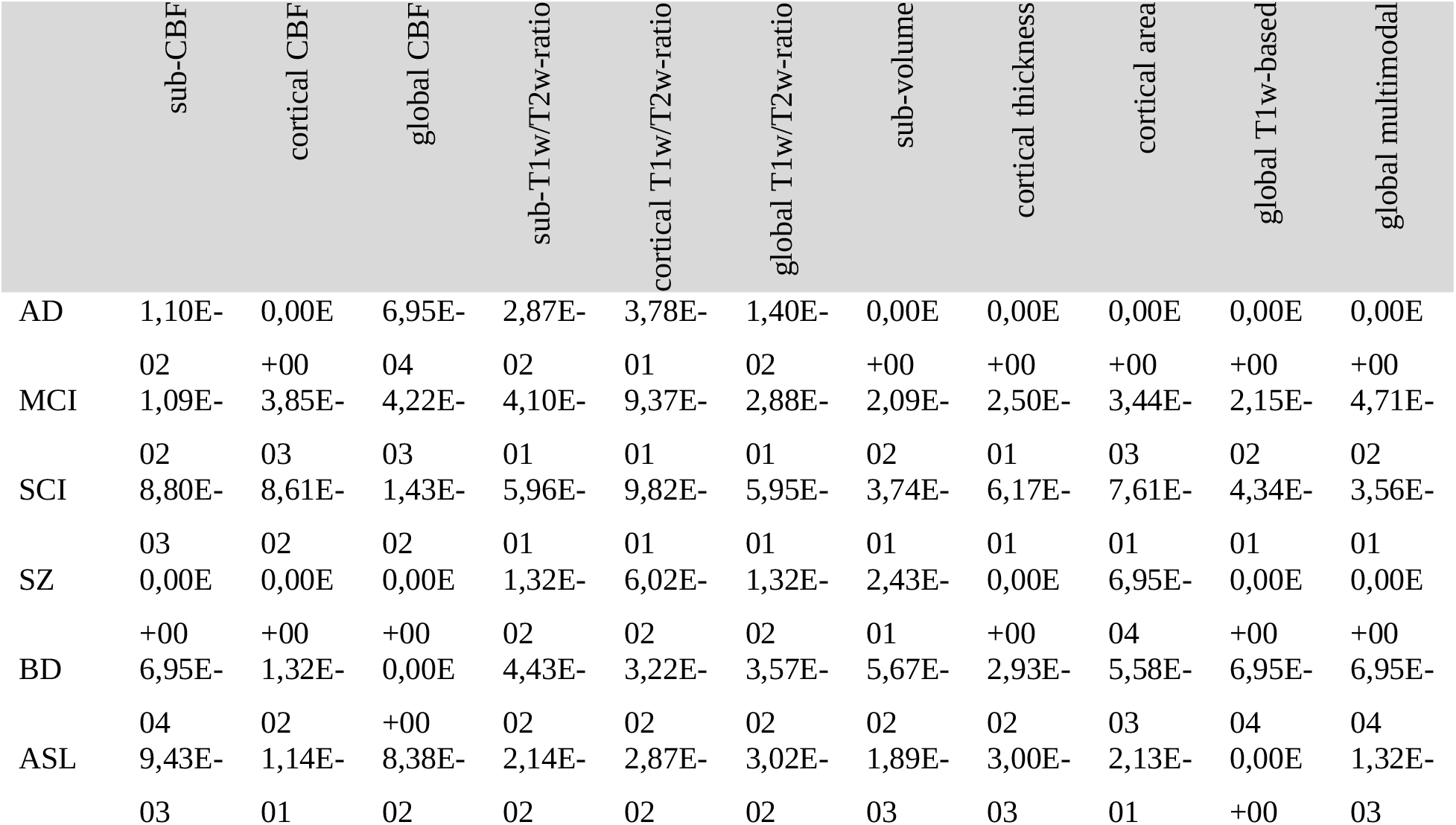
p-values associated with each AUC, after performing 5000 permutations.

